# Predicting survival and trial outcome in non-small cell lung cancer integrating tumor and blood markers kinetics with machine learning

**DOI:** 10.1101/2023.09.26.23296135

**Authors:** Sébastien Benzekry, Mélanie Karlsen, Célestin Bigarré, Abdessamad El Kaoutari, Bruno Gomes, Martin Stern, Ales Neubert, Rene Bruno, François Mercier, Suresh Vatakuti, Peter Curle, Candice Jamois

**Affiliations:** COMPutational pharmacology and clinical Oncology Department, Inria Sophia Antipolis–Méditerranée, Cancer Research Center of Marseille, Inserm UMR1068, CNRS UMR7258, Aix Marseille University UM105, Marseille, France; Pharma Research and Early Development, Early Development Oncology, Roche Innovation Center Basel, Switzerland; Pharma Research and Early Development, Early Development Oncology, Roche Innovation Center Zurich, Switzerland; Pharma Research and Early Development, Data & Analytics, Roche Innovation Center Basel, Switzerland; Modeling and Simulation, Clinical Pharmacology, Genentech Research and Early Development, Marseille France; Modeling and Simulation, Clinical Pharmacology, Genentech Research and Early Development, Roche Innovation Center Basel; Pharma Research and Early Development, Predictive Modeling and Data Analytics, Roche Innovation Center Basel, Switzerland; Inovigate, Basel, Switzerland; Pharma Research and Early Development, Translational PKPD and Clinical Pharmacology, Roche Innovation Center Basel, Switzerland

## Abstract

Existing survival prediction models rely only on baseline or tumor kinetics data and lack machine learning integration. We introduce a novel kinetics-machine learning (kML) model that integrates baseline markers, tumor kinetics and four on-treatment simple blood markers (albumin, CRP, lactate dehydrogenase and neutrophils). Developed for immune-checkpoint inhibition (ICI) in non-small cell lung cancer on three phase 2 trials (533 patients), kML was validated on the two arms of a phase 3 trial (ICI and chemotherapy, 377 and 354 patients). It outperformed the current state-of-the-art for individual predictions with a test set c-index of 0.790, a 12-months survival accuracy of 78.7% and a hazard ratio of 25.2 (95% CI: 10.4 – 61.3, *p* < 0.0001) to identify long-term survivors. Critically, kML predicted the success of the phase 3 trial using only 25 weeks of on-study data (predicted HR = 0.814 (0.64 – 0.994) versus final study HR = 0.778 (0.65 – 0.931)). Our model constitutes a valuable approach to support personalized medicine and drug development.

## Introduction

Lung cancer is the leading cause of cancer death worldwide^1^, with non-small cell lung cancer (NSCLC) being the most prevalent type, representing 80% – 85% of case^2^. Immune-checkpoint inhibitors (ICI) (e.g., atezolizumab (ATZ)) have led to significant improvements in survival rates for patients with advanced cancers such as NSCLC^3,4^. However, there is still a large variability in clinical response and progression eventually occurs in a majority of patients^5^. Additionally, drug development in immuno-oncology is highly challenging, with a 95% attrition rate^6^. Current approaches for go/no-go decisions are based on interim endpoints (e.g., progression-free survival, overall response rate) that have often been found to be poor predictors of the primary endpoint of most clinical trials in oncology, overall survival (OS)^7^. This calls for better surrogate markers at interim analyses. Altogether, there is a need for better and validated predictive models of OS for both personalized health care (individual predictions) and drug development (trial predictions).

Currently, PDL1 expression is the only routine biomarker used for NSCLC patients^5,8^ despite being controversial^9,10^. Tumor mutational burden^8,11,12^ and transcriptomic data^5,13,14^ have also been investigated but did not reach clinical practice. Here we posit that such static and single marker approach is intrinsically limited and that substantial additional predictive performances could be gained by: 1) using multi-modal integrative analyses relying on a combination of markers and machine learning algorithms^5,12,14,15^ and 2) including dynamic markers obtained from early on-treatment data^15,16^. The nonlinear mixed-effects (NLME) modeling approach is well suited for the latter^17^, and tumor kinetics (TK) model-based metrics have been shown to carry significant predictive value for OS in oncology, including ATZ monotherapy in advanced NSCLC^18–20^. The first main novelty of the current study is to establish the predictive value of model-based parameters of simple blood markers kinetics (BK), in addition to TK.

The second main novelty is to apply machine learning (ML) algorithms, increasingly used in biology and medicine^21^ but only rarely for TK-OS modeling^22^, instead of classical survival models. Extensions of classical ML models to survival data have been proposed (e.g., random survival forests^23^), but their actual superiority over standard approaches remains controversial^24^. In addition, most ML studies to date are underpowered due to low sample sizes in both training and test sets.

Here, we coupled the strengths of NLME modeling with ML to derive a predictive model of OS from baseline and on-treatment data, called kinetics-machine learning (kML, Figure 1A). We leveraged large training and test datasets to achieve robust results (Figure 1B). Subsequently, we tested the operational predictive capabilities of kML in two relevant scenarios: 1) individual prediction of OS and 2) prediction of the outcome of a phase 3 trial from early on-study data.

**Figure 1.**
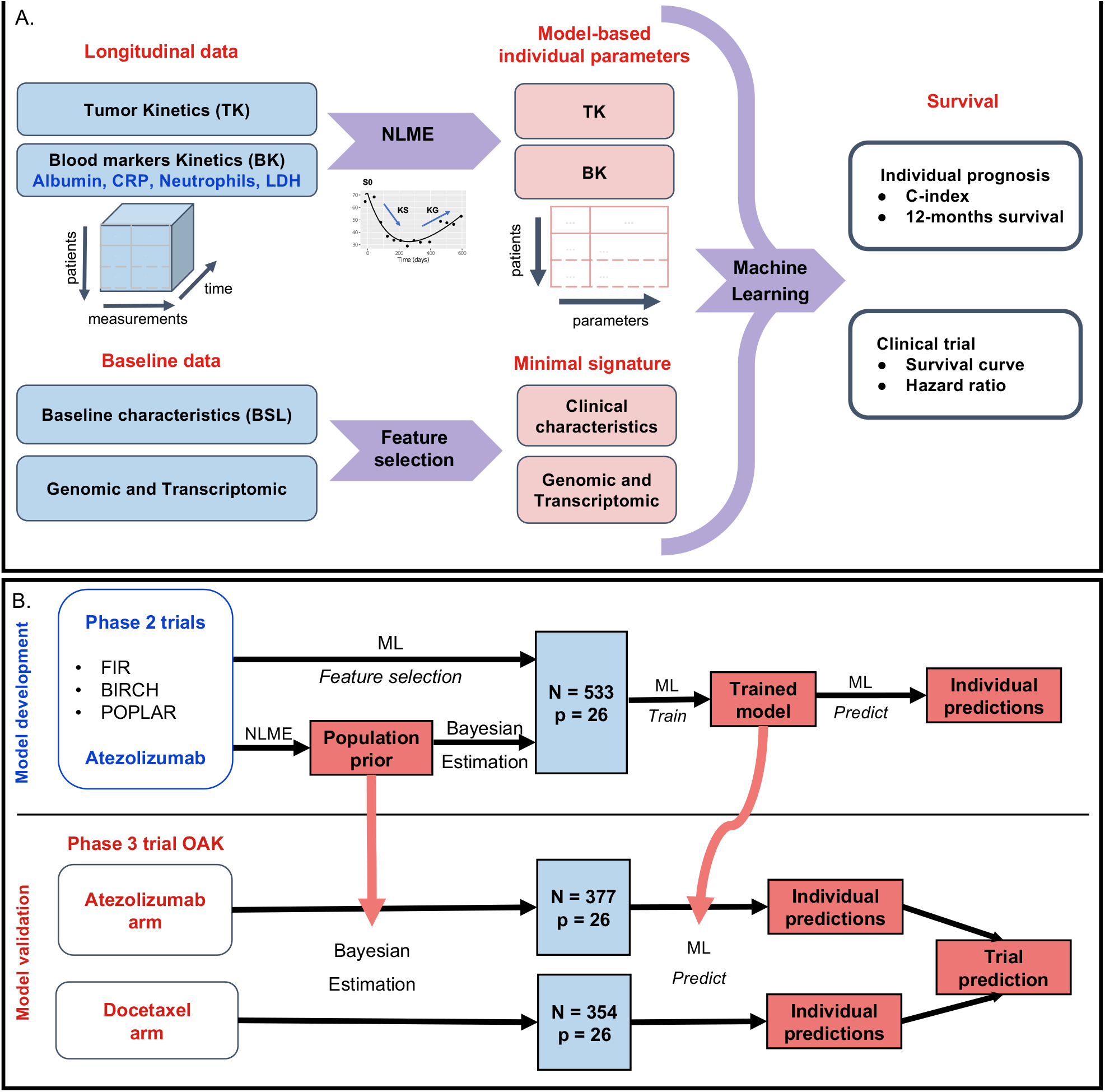
Study schematic. **A.** Baseline and longitudinal data were combined into a machine learning algorithm in order to predict individual survival prognosis. Longitudinal data were modelled using nonlinear mixed-effects modelling, whereas machine learning-based feature selection was applied to the baseline data to derive a minimal signature. Tumor kinetics and biological kinetics parameters were combined with the minimal signature to predict survival. Predictive performances were assessed using survival metrics (c-index and survival at horizon times). **B.** Algorithm used to develop the model on the train data and carry it to the test set for external validation. Each step — preprocess, learning of the Bayesian priors, dimensionality reduction, feature selection, choice, tuning and training of the machine learning algorithm — were calibrated on the training set and then applied to the test set. TK: tumor kinetics; BK: blood markers kinetics; ML: machine learning; NLME: nonlinear mixed-effects modelling

## Methods

### Data

For both training and external validation (testing) sets, patients from French centers were excluded for legal reasons (*N* = 118, not included in the numbers above). The training set comprised the FIR (NCT01846416)^25^, POPLAR (NCT01903993)^3^ and BIRCH (NCT02031458)^26^ phase 2 clinical trials. The test set was the atezolizumab arm of the OAK phase 3 trial (NCT02008227)^27^ for individual predictions and additionally the docetaxel arm for trial predictions (Supplementary Figure 1). These studies were conducted in accordance with the Declaration of Helsinki after approval by institutional review boards or independent ethics committees. All patients provided written informed consent.

The outcome considered was overall survival (OS), defined as the time between treatment start and death or last follow-up, in which case the data was right-censored. The median follow-up was 35.2 months (95%CI:34.5–35.7) in the training set and 26.8 months (95%CI:26.3–27.5) in the test set.

### Preprocessing

#### Baseline data

The baseline data consisted of 63 variables spanning demographic and biological data, clinical information and disease status (see Supplementary Figure 2–4 for a description of the main variables). PD-L1 expression on tumor cells was measured by immunohistochemistry or quantitative polymerase chain reaction, with four possible levels (0: < 1%; 1: ≥ 1%; 2: ≥ 5% and 3: ≥ 50%)^3^. We refer to the above-mentioned identifiers and references for further details on the other variables. Data were measured in accordance to the studies principles.

#### Tumor and blood markers kinetics (TK and BK)

Patients with only one baseline SLD measurement and no SLD measurement during the treatment period were excluded (*N* = 110). For BK, first time points prior to treatment start were discarded. Then, four exclusion rules were established to identify anomalous data points: 1) values outside physiologically possible bounds, 2) duplicates, 3) values that abruptly went to an extreme out-of-range value between two measurements, 4) only the BK value at the closest time point to treatment initiation was kept. Eventually, in order to have sufficient data for Bayesian estimation with early data, patients with less than three observations before cycle 5 were removed. We refer to the supplementary methods for details.

### Nonlinear mixed-effects modeling

#### Population approach

Statistical hierarchical nonlinear mixed-effects modeling (NLME) was used to implement a population approach^28^ for the kinetic data and parameter estimation was conducted using the Monolix software^29^ Mathematical details are given in the supplementary methods.

#### Structural models

Following previous work, the TK structural model was assumed to be the sum of two exponentials^19,30^:

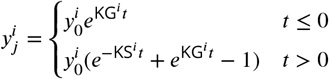

where *t* = 0 corresponds to treatment initiation and *y*_0_, KG and KS are three parameters, representing respectively the baseline value, growth and shrinkage rates. This model was also considered for BK, together with three other models: constant 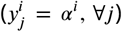, linear 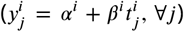 and hyperbolic 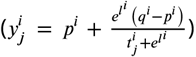^31^. Quantitative comparison of goodness-of-fit between models was assessed using the corrected Bayesian information criterion^32^.

#### Identification of individual model-based parameters

The population parameters identified on the training set were used to define prior distributions of the TK and BK model parameters. These “training” priors were used for Bayesian estimation (maximum a posteriori estimate) of the individual TK and BK model parameters, not only for the training set but also for the test sets, in order to avoid leakage. To focus on the pure kinetic parameters, the model-estimated baseline parameters were not kept. We additionally considered the ratio of the model-predicted value at cycle 3 day 1 to the model-estimated baseline parameter. Altogether, there were three individual parameters for each marker: X_*KG*_, X_*KS*_ and X_*ratio*_ for *X* = TK, CRP, LDH and neutrophils; and albumin_*p*_, albumin_*l*_ and albumin_*ratio*_ for albumin.

#### Truncated data: individual-level

Individual-level truncated datasets were derived from the longitudinal TK and BK data by keeping data only up to: cycle 3 day 1 (C3D1, 1.5 months), C5D1 (3 months) and C10D1 (6.75 months). New training priors were estimated from each CXD1 training set. The resulting TK and BK truncated model parameter Y for marker *X* at cycle *i* were denoted by *X*_Y, *i*_ (e.g., *ldh*_KG, 5_).

#### Truncated data: study-level for trial prediction

Study-level truncated datasets were defined at the following on-study landmark times *lt* after study initiation (first patient recruited): *lt* = 10, 25 and 60 weeks.

Only the patients enrolled before this time and their data collected up to *lt* was used. Note that here *t* = 0 corresponds to study initiation and thus patients in these datasets have varying follow-up duration (from 0 to *lt*), in contrast to individual-level truncated datasets.

### Machine learning

#### Data preparation

Missing values (1.6% total, maximum 12% in one variable) were imputed with the median for numeric variables and mode for categorical variables, learned on the training set, even when applied to the test set. All numeric variables were centered and scaled. Means and standard deviations were learned on the train and carried to the test set.

#### Models

Model elaboration and development was performed exclusively on the training set, using 10 folds cross-validation for predictive performances evaluation. Due to censoring in the data, survival models were used: proportional hazards Cox regression^33^, extreme gradient boosting (XGB) with either Cox or accelerated failure time (AFT) models^34^ and random survival forests (RSF)^23^. Nested cross-validation with inner bagging in each 10-fold cross-validation outer loop was used to evaluate the benefit of tuning the hyperparameters^35^. Improvement of the performances was negligible with hyperparameter tuning (Supplementary Figure 5). Therefore, we used the default values of the hyperparameters. For the final RSF model: number of trees *ntree* = 500, number of variables to possibly split at each node *mtry* = 5, minumum size of terminal node *nodesize* = 15, number of random splits for splitting a variable *nsplit* = 10.

#### Evaluation

Predictive performances were assessed for either discrimination (c-index and classification metrics at horizon times τ), calibration (calibration curves) or stratification (dichotomized KM survival curves). For each individual, the RSF model gives two prediction outputs: a scalar value termed “mortality” that we will refer to as “ML score”, and time-dependent predicted survival curves^23^. The former was used to compute the c-index using the `rcorr.cens` function of the `hmisc` R package^36,37^. For prediction of survival at a horizon time τ, we used the latter to compute model-predicted probabilities of death at τ. Unless otherwise specified, τ = 12 months. Survival-adapted metrics of predictive performance were used for sensitivity, specificity, area under the receiver-operator curve (ROC AUC) and negative and positive predictive value (NPV and PPV) to account for censoring^38,39^. For computation of accuracy, censored patients before τ were discarded (*N* = 17/396 in the test set at 12 months). The optimal cut-points used for individual OS predictions on the test set were defined as the Kaplan-Meier estimated survival probability in the training set at τ (0.257 at 6 months, 0.437 at 12 months, 0.634 at 24 months).

For patient stratification (dichotomized KM curves), the ML score was used, with models trained on the training set and predicted on the test set. In order to assess stratification abilities to capture the 20% of long-term survivors, cut-points were set at the 20^th^ percentiles for each variable/score evaluated. This cut-point arbitrary definition was also motivated by the aim to ensure fair comparison between multiple parameters on the same data. Significance of differences in KM curves was established using the logrank test, and hazard ratios were computed using proportional hazards Cox regression.

#### Variable selection and minimal signature

Variable selection was performed only for the BSL data. The method was based on two steps: 1) sorting the variables using least absolute shrinkage and selection operator (LASSO)^40^ and 2) building RSF incremental models including increasing numbers of variables. LASSO sorting was defined as taking the coefficients gradually becoming non-zero during likelihood maximization when the regularization parameter decreases. The minimal signature was defined as the minimal set of variables able to achieve a c-index larger than 0.75 and an AUC larger than 0.8, with the addition of 4 well-established prognosis.

#### Survival simulations and computation of predicted HRs

For each patient *i*, one output of the kML model is a survival curve *S*^*i*^ (*t*). This gives the cumulative distribution function 1 − *S*^*i*^ (*t*) of the random variable *T* ^*i*^ of the time to death for patient *i*, which was used to simulate 100 replicates of *T* ^*i*^. Pooling all patients together, we thus obtained 100 replicates of {*T* ^*i, AT Z*^, *T* ^*j, DT X*^} for *i* and *j* being the patient indices within the ATZ and docetaxel arms, respectively. Each replicate then led to 1) a predicted survival curve in each arm and 2) a Cox proportional hazard HR between the two arms. Taking the mean and the 5^th^ and 95^th^ percentiles over all replicates yielded the reported point estimate and corresponding 95% prediction interval. The same procedure was used for study-truncated data.

## Data Availability

Qualified researchers may request access to individual patient level data through the clinical study data request platform (https://vivli.org/). Further details on Roche’s criteria for eligible studies are available here (https://vivli.org/members/ourmembers/). For further details on Roche’s Global Policy on the Sharing of Clinical Information and how to request access to related clinical study documents, see here https://www.roche.com/innovation/process/clinical-trials/data-sharing/.

## Code availability

Algorithms used for data analysis are all publicly available from the indicated libraries and references in the Methods section.

## Results

### Data

The data consisted of advanced NSCLC patients enrolled in ATZ trials (N = 1936, Figure 1B and Supplementary Figure 1). Three ATZ phase 2 trials were pooled into a training dataset^3,25,26^ (*N* = 862). The external validation (test) set comprised data from the ATZ arm (*N* = 553) of the OAK phase 3 trial^27^. For trial outcome prediction the docetaxel arm (*N* = 521) was added as an additional test set.

Variables comprised baseline (pre-treatment) and longitudinal (on-treatment) data (Figure 1A). The former included: patients and disease characteristics (*p* = 63 variables, 43 numeric and 20 categorical, denoted BSL) and transcriptomic (“RNAseq”, *p* = 58, 311 transcripts) data. The latter included: longitudinal investigator-assessed sum of largest diameters (SLD) of lesions as per the RECIST criteria^41^, denoted by tumor kinetics (TK, *k* = 5, 473/3, 015 time points in the train/test sets, respectively, median 5/4 data points per patient, range 2/2 —24/20); and longitudinal measurements of four blood markers (albumin, C-reactive protein (CRP), lactate dehydrogenase (LDH) and neutrophils), denoted together as blood markers kinetics (BK, *k* = 60, 779/38, 460 data points, median 11–7–11–11/9–9–9–10 data points per patient, range 3–3–3–3/3–3–3–3 —60–63–63–78/82–47–77–89 for albumin–CRP–LDH-neutrophils in the train/test sets, respectively).

### Nonlinear mixed-effects modeling (NLME) of longitudinal markers

We first developed NLME models for the longitudinal data (Figure 1B). The TK structural model was the sum of an increasing and a decreasing exponential function (double exponential model)^30^. It was able to accurately describe the training data with no goodness-of-fit misspecification (Figure 2A and Supplementary Figure 6). Population parameters were estimated with good accuracy (all relative standard errors smaller than 9%, Table 1).

**Table 1.**
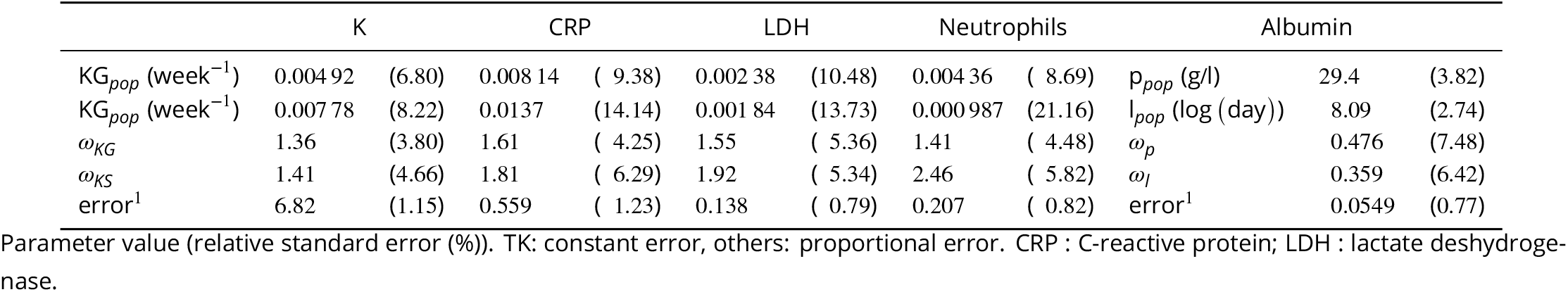
Parameters from nonlinear mixed-effects modeling of tumor and blood marker kinetics.

**Figure 2.**
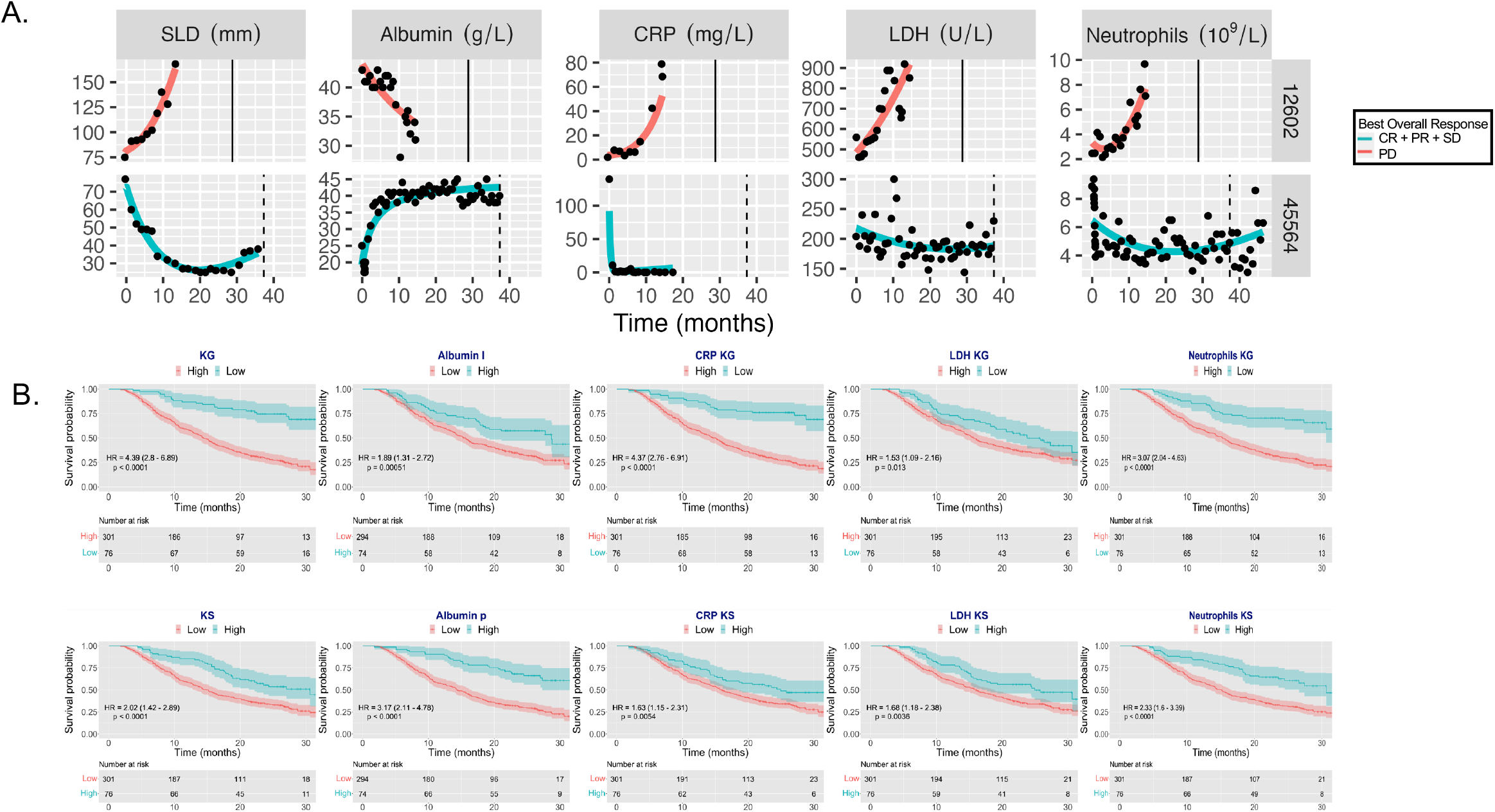
Goodness-of-fit metrics and plots of dynamic BK models. **A.** Representative individual fits for the TK and BK best empirical models showing non-trivial kinetic parameters well captured by the dynamic models. Survival is indicated by a vertical line (solid = death, dashed = censored). **B.** Stratified Kaplan-Meier curves at the 20^th^ percentile level on the test set, for TK and BK model-based parameters. Missing values were removed in this univariable analysis, explaining the difference of initial number of patients for albumin that had 9 patients in this case. CR: complete response; PR: partial response; SD: stable disease; PD: progressive disease.

To analyze the BK data, we first investigated whether significant kinetic patterns could be observed beyond random noise (due to, e.g., measurement errors, see raw data in Supplementary Figures 7–10). The latter was considered as the null hypothesis, described by a constant model. It was tested against three alternative empiric models: linear, hyperbolic (monotonous but non-linear and saturating) and double-exponential (nonlinear and non-monotonous). For all four BKs, we found significant kinetics compared with the constant model, as shown by lower corrected Bayesian information criterion and relative error between model fits and data (Supplementary Figure 11). The best descriptive models were hyperbolic for albumin and double-exponential for the other BKs. Individual fits to patient kinetics with the best models showed substantial descriptive power (Figure 2A), which was confirmed by data versus model fits plots (Supplementary Figures 12–15). Parametric identifiability of population parameters was excellent for all models (Table 1).

We further assessed the stratification value of the individual model-based kinetic marker for OS prognosis (Figure 2B). The TK parameter KG (growth rate) exhibited good stratifying ability (HR = 4.39 (2.8 – 6.89)), which was similar to the CRP_*KG*_ parameter (HR = 4.37 (2.76 – 6.91)). Ranked by HR importance; (controlled by the 20^th^ percentile definition of the cut-point, see methods), the following four best parameters were albumin_*p*_ (HR = 3.17 (2.11 – 4.78)), neutrophils_*KG*_ (HR = 3.07 (2.04 – 4.63)), neutrophils_*KS*_ (HR = 2.33 (1.6 – 3.39)) and TK_*KS*_ (HR = 2.02 (1.42 – 2.89)). All kinetic parameters carried substantial prognostic power (*p* < 0.0001, log rank test).

For TK and BKs we complemented the initial model parameters with an additional metric that was considered valuable for early prediction: the model-predicted ratio of change over baseline at cycle 3 day 1.

### Survival prediction using kinetics-machine learning (kML): model development

Four feature sets resulted from the analysis above: BSL, RNAseq, TK and BK (Figure 1A). The development of a kinetics-machine learning (kML) comprised two main steps: choice of the algorithm and derivation of a minimal signature (Figure 1B). The first was achieved by benchmarking four models that used all variables (*p* = 119, *N* = 553). The random survival forest (RSF) model found to exhibit the best performances (Supplementary Figure 5) and was thus selected. Notably, we found significantly better predictive performances of RSF over a classical Cox proportional hazard regression model (*p* = 0.0006).

Feature selection on BSL variables was performed building incremental RSF models based on LASSO importance-sorted variables (Figure 3A). The model using all of them achieved the best score. Nevertheless, keeping in mind the objective to ultimately support decision making and patient stratification, a minimal (11 features), near-optimal, set of BSL variables was selected and denoted mBSL. It was defined as the first seven variables reaching the plateau (CRP, heart rate, neutrophils to lymphocytes ratio, neutrophils, lymphocytes to leukocytes ratio, liver metastases and ECOG score), complemented with four variables with established prognostic or predictive value and available in routine care: PD-L1 expression (50% cut-off)^3^, hemoglobin^42^, SLD^22^ and LDH^43,44^.

**Figure 3.**
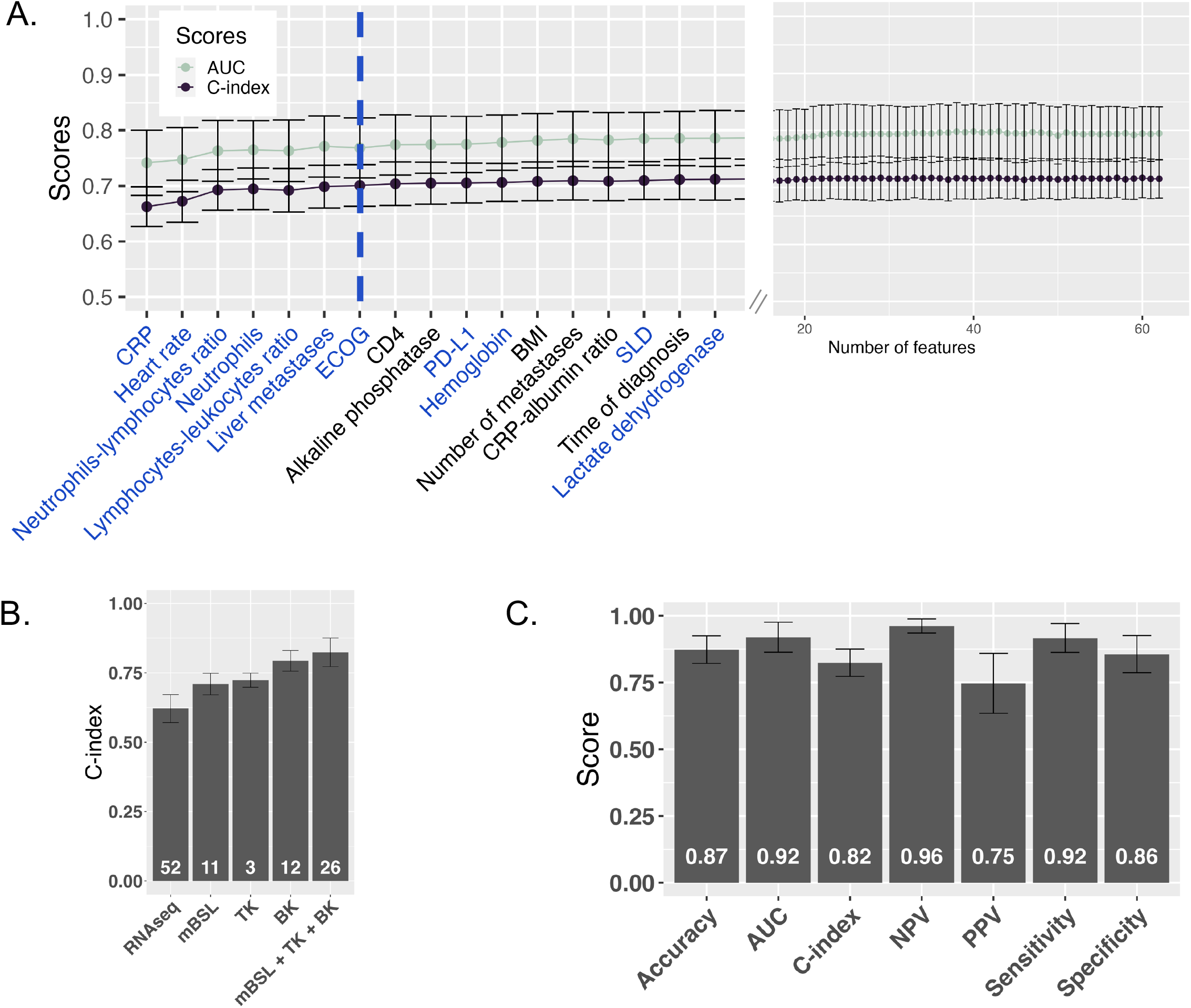
Minimal baseline (mBSL) signature and kinetics-ML (kML) model. **A.** Cross-validated (CV) performance scores on the training set (c-index and AUC, mean ± standard deviation) for incremental random survival forest (RSF) models using an increasing number of baseline clinical and biological variables sorted by LASSO importance. The dashed blue line shows the minimal number of variables reaching the plateau. Blue-colored variables correspond to the minimal clinical signature (mBSL). **B.**Comparative CV c-indices of RSF models based either on RNAseq, mBSL, TK, BK and mBSL + TK + BK (final model, kML) variables showing increased predictive performances over baseline when using model-based parameters of kinetic markers. Numbers on the bars indicate the number of variables. **C.** CV performances of the kML model for discrimination (c-index) and classification (survival prediction at 12-months OS).

Applying stringent criteria to the RNAseq data (see supplementary methods), we selected 167 transcripts as candidates for final variable selection using Bolasso regression model to identify the optimal set of predictors^45^. Finally, we ended up with 52 RNAseq variables that corresponded to the highest average c-index of 0.64.

We then compared the cross-validated c-index of each feature set on the train data (Figure 3B). Because of negligible discrimination performances (*c-index* = 0.62 ± 0.050) and non-systematic availability of those data, the RNAseq set was removed from the model. The selected set of clinical data at baseline (mBSL) exhibited moderate discrimination performances (*c-index* = 0.710 ± 0.038), which was slightly outperformed by the TK set (*c-index* = 0.723 ± 0.025). Interestingly, the BK set significantly outperformed both baseline clinical and TK (*c-index* = 0.793±0.038, *p* = 0.0004 and 0.0005 respectively, Student’s t-test). Jointly, mBSL, TK and BK performed significantly better than any feature set alone (*c-index* = 0.824 ± 0.050, *p* = 0.00007, 0.0002 and 0.055), as well as any combination of two sets among the three (mBSL + TK: *c-index* = 0.77 ± 0.026, mBSL + BK: *c-index* = 0.81 ± 0.027, TK + BK: *c-index* = 0.80 ± 0.049). The resulting model combining mBSL, TK and BK was denoted kML (kinetics-machine learning).

During cross-validation on the training set, kML exhibited excellent predictive performances across multiple metrics, with minimal between-folds variability (*AUC* = 0.919 ± 0.056, *accuracy* = 0.873 ± 0.052, Figure 3C).

#### External validation

The predictive performance of the final kML model (mBSL, TK and BK) was assessed on the ATZ test set (377 patients). At the population level, the model-predicted survival curve was in excellent agreement with the observed data (Figure 4A). Notably, the prediction interval from the model was narrow, indicating high precision. At the individual level, consistent with the cross-validation results, substantial discrimination performances were observed (*c-index* = 0.790, accuracy and AUC for 12-months survival probability 0.787 and 0.874, respectively, Figure 4B). All classification metrics for prediction of survival at 12 months were high (≥ 0.78), except PPV, indicating worse ability to predict death than survival. Although smaller, they were similar to the cross-validation results.

**Figure 4.**
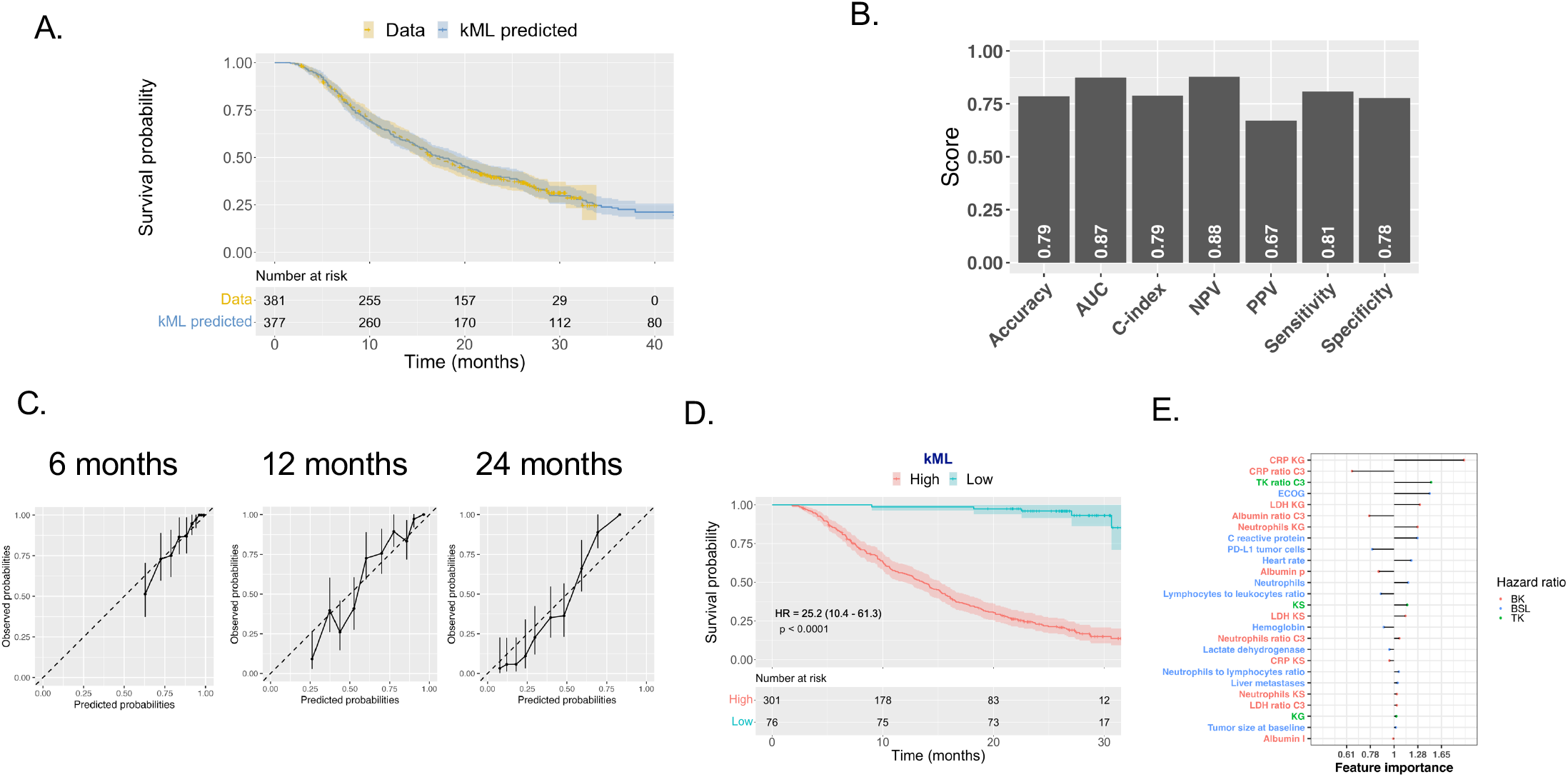
Predictive performances of kML on the ATZ test set. **A.** Comparison of the population-level survival curves between the data (KM estimator) and the model prediction. **B.** Scores of discrimination metrics. Classification metrics were computed for prediction of OS at 12 months. **C.** Calibration curves at 6, 12 and 24 months, showing the observed survival probabilities (with KM 95% confidence interval) versus the predicted ones in 10 bins corresponding to the model-predicted survival probability deciles. Dashed line is the identity. **D.** Dichotomized KM survival curves based on the ML model-predicted score (high versus low), at the 20^th^ percentile cut-off. **E.** Variables importance (multivariable hazard ratios) in the full time-course kML model.

In addition, calibration curves revealed good performance, at multiple horizon times (Figure 4C). Model-predicted probabilities were concordant with the observed KM estimates of the survival probabilities, over the entire range of the binned predicted probabilities. This is further illustrated by the contingency Table 2. For instance, among 212 patients predicted to be alive at 12 months, 182 (85.8%) were actually alive. Predictive AUC was good at other horizon times (0.846 and 0.910 at 6 and 24 months, respectively, Supplementary Figure 16). However, PPV and sensitivity were very low at 6 months.

**Table 2.** Contingency table for OS prediction at 12 months.

|  |  | TRUTH |  |  |  |  |
| --- | --- | --- | --- | --- | --- | --- |
|  |  | Alive (0) |  | dead (1) |  | Total |
| MODEL | Alive (-) | 182 |  | 30 |  | 212 (58.7%) |
|  | Dead (+) | 48 |  | 101 |  | 149 (41.3%) |
|  | Total | 230 | (63.7%) | 131 | (36.3%) | 361 |

Note: 16/377 censored patients with survival time $\leq 12$ months removed for computation of accuracy. sensitivity, specificity, PPV and NPV don't correspond exactly to the numbers because they are computed from KM estimate, thus adjusting for censoring bias.

Notably, the kML mortality score derived from the model and learned on the training set was able to accurately stratify OS in the test set (HR = 25.2 (10.4 – 61.3), *p* < 0.0001, Figure 4D), indicating excellent ability to identify the 20% of long-term survivors. It outperformed all single kinetic markers (Figure 2C).

Variables importance was assessed by running a post-hoc multivariable Cox regression (Figure 4F). Interestingly, the top two variables were BKs (CRP_*KG*_ and CRP ratio C3). In addition, TK and BK made up for six out of the seven top important features and were found more important than PD-L1.

Given the large sample size of our data, we further assessed the model performances when trained on smaller data sets (Supplementary Figure 17). The learning curve revealed that approximately 200 patients were necessary to reach similar performance to the ones obtained with the full training set (*N* = 533), for both cross-validation and external validation on the test set (*c-index* = 0.82 ± 0.056 vs *c-index* = 0.82 ± 0.050 in cross-validation, 0.78 vs 0.79 on the test set, models trained with 200 vs 533 patients, respectively). Trained with only 60 patients, kML reached already good performances (*c-index* = 0.76 ± 0.15 and 0.74 in cross-validation and test, respectively).

Together, these results demonstrate important predictive performances of overall survival following ATZ treatment using the kML model.

### Application to individual survival prognosis from early on-treatment data

Results above required full on-treatment time-course data to compute TK and BK markers, thus cannot be used to make early predictions. To investigate the operational applicability of our methodology, data from the test set were truncated at the beginning of treatment cycles number 3, 5 and 10, respectively corresponding to 1.5, 3 and 6.75 months. We found that integrating longer on-treatment data in kML, the predictive performances steadily increased (Figure 5A and Supplementary Figure 18). Using the baseline variables only (mBSL), the stratification ability was significant but moderate (HR = 1.74 (1.24 – 2.46), *p* = 0.0014, Figure 5B). In contrast, kML exhibited increasing stratification ability from data at 1.5 months (HR = 2.19 (1.53 – 3.12), *p* < 0.0001), 3 months (HR = 3.51 (2.33 – 5.3), *p* < 0.0001) and 6.8 months (HR = 5.01 (3.16 – 7.95), *p* < 0.0001), see Figure 5C.

**Figure 5.**
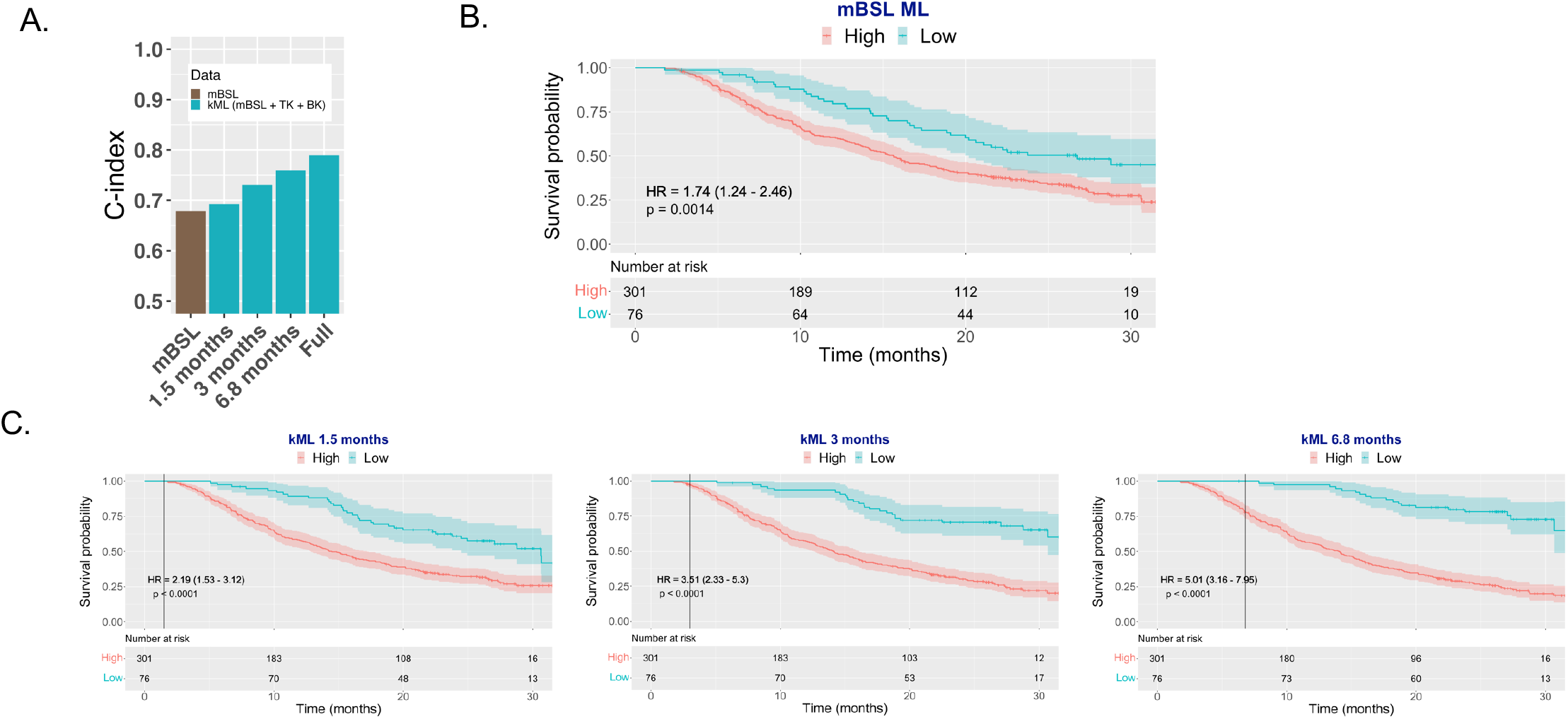
Predictive value of kML from cycle-truncated data. **A.** Predictive power (c-index) of ML models using baseline (BSL) or truncated data at 1.5, 3 and 6.8 months as well as the full time-course. **B.** Stratified KM survival curves using a RSF model trained on the minimal baseline (mBSL) variables. **C.** Stratified KM survival curves using kML from 1.5 months (2 cycles), 3 months (4 cycles) and 6.8 months (9 cycles) truncated data. Truncation time is indicated by the vertical line. TK: tumor kinetics; BK: biological kinetics; LDH: lactate dehydrogenase; CRP: C-reactive protein.

Further investigation of the predictive performances of individual kinetic markers revealed that TK parameters were the most informative at 6 weeks (1.5 months, first imaging assessment). Adding BKs to TKs brought additional predictive value starting at 3 months, and BKs outperformed TK from 6.75 months on (Supplementary Figure 19A). Among BKs, neutrophils kinetics appeared to be the most predictive, followed by CRP, albumin and LDH. However, the combined BK signature outperformed each individual BK, indicating that their collective predictive capabilities were not driven by any single biomarker alone.

Interestingly, the most important variable at 1.5 months was a kinetic one, TK ratio C3 with following variables being from mBSL (e.g., liver metastases, PDL1 and ECOG). When more on-treatment variables become available, this shifted to TK and BK (TK ratio C3, TK_*KS*_, TK_*KG*_, CRP_*KG*_, LDH_*KG*_), see Supplementary Figure 19B.

### Application to clinical trial outcome prediction from early on-study data

The kML model can also be applied for the prediction of the outcome of a clinical trial (survival curves and associated hazard ratio), from early on-study data. We performed on-study runcations on the test set based on a number of weeks after the date of the first patient recruited (see methods). Here, we applied the model to predict not only patients receiving ATZ, but also docetaxel (Figure 1B). Predictions of the kML model applied to each arm yielded very accurate results when using data from the entire study (predicted HR = 0.784 (0.7 – 0.842)), versus data HR = 0.778 (0.65 – 0.931), Figure 6A–B). Notably, the model prediction intervals were narrower than the data Kaplan-Meier confidence intervals, probably because the kML-trained model incorporates the information from the three phase 2 trials. Using only early data, the model was already able to detect a (non-significant) tendency at 10 weeks, with only 23 and 30 patients in each arm, and very short follow-up. Starting from data available at 25 weeks (6.25 months), the model correctly predicted a positive outcome of the study, with a 95% prediction interval of the HR below 1. Of note, the available data at this time (dashed lines, Figure 6A and red HR CIs in Figure 6B) was far from being conclusive. The model prediction was stable from 25 weeks on whereas the OS data only exhibited significant HR starting from 60 weeks and required more than 300 patients in each arm to be conclusive.

**Figure 6.**
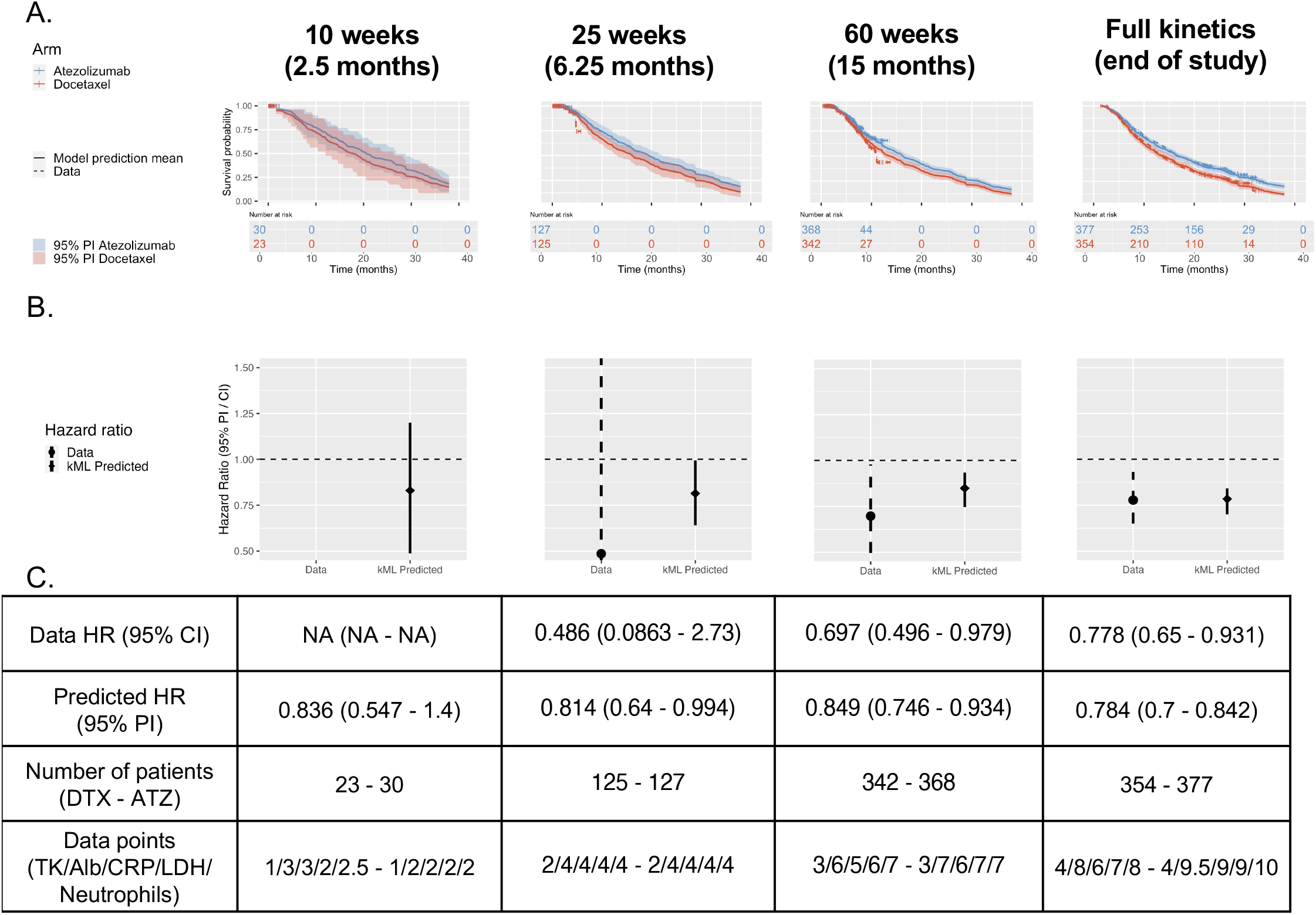
Use of kML for early-prediction of the outcome of a clinical trial. **A.** Survival curves model-based predictions and prediction intervals versus actual data from on-study data at multiple horizon times after study initiation. Note that the model is able to predict full survival curves even if based on early kinetics. **B.** Compared data and kML-predicted hazard ratios. **C.** Description of hazard ratios, number of patients and number of data points available in each arm, at the landmark on-study time points. PI: prediction interval, CI: confidence interval, DTX: docetaxel arm, ATZ: atezolizumab arm.

## Discussion

Blood markers from hematology and biochemistry are routinely collected during clinical care or drug trials. They are cost-effective and easily obtained both before and during treatment. There is limited exploration regarding the predictive capabilities of the kinetics of such data. Combining BSL variables with on-treatment data (TK and BK), we addressed this question using a novel hybrid NLME–ML methodology. The resulted kML model demonstrated excellent predictive performances for OS in two aspects: 1) patient-level predictions (discrimination, calibration and patient stratification) and 2) trial-level predictions. The kML model outperformed current state-of-the-art methods based on either baseline or on-treatment data alone, utilizing only routine clinical information, with a c-index of 0.79 and an accuracy of 78% for prediction of 12-month survival, on the test dataset. Overall, kML incorporates 26 features, out of which 15 features require monitoring five quantities over time (tumor size, albumin, CRP, LDH and neutrophils).

Regarding baseline markers, the predictive value of PD-L1 expression, commonly used in clinical care, is controversial^9,10^. Previous studies reported an AUC for durable response of 0.601 and a PFS HR of 1.90 (PD-L1 ≥ 1% vs 0%)^8^. Baseline tumor mutational burden showed similar predictive value initially (*AUC* = 0.646)^11^, but led to disappointing results in a recent prospective study^46^. Baseline blood counts were previously reported to predict overall survival^43,47–49^ and treatment response (AUC = 0.74)^42^. The ROPRO score, derived from a large pan-cancer cohort and incorporating baseline clinical and biological data (27 variables) achieved a c-index of 0.69 and a 3-months AUC of 0.743 for prediction of survival in the OAK clinical trial^50^. Here, we confirmed these findings and established a minimal signature of such data composed of only 11 variables (CRP, heart rate, neutrophils to lymphocytes ratio, neutrophils, lymphocytes to leukocytes ratio, liver metastases, ECOG, PD-L1 ≥ 50%, hemoglobin, SLD and LDH), yet with similar predictive performances (*c-index* = 0.678) and significant stratification ability (*HR* = 1.74, *p* = 0.0014). Altogether, our kML model demonstrated substantially better predictive performances than these baseline models.

We further confirmed the established predictive value of TK model-based parameters^19,20^. Blood- or serum-derived longitudinal markers kinetics have to date rarely been modeled. Gavrilov et al. proposed to model NLR kinetics and demonstrated improved OS predictions over TK alone^31^. Here we extended to four BKs: albumin, CRP, LDH and neutrophils. This choice was not only motivated by observed statistical associations, but also from biological considerations. Albumin is associated with nutritional status (cachexic state) and is known to evolve with time in responders. CRP is a marker of systemic inflammation^44^. Increased CRP, decreased albumin level, and increased CRP/albumin ratio have been reported to be associated with poor survival^51^. Neutrophils play a role in inflammation by promoting a favorable microenvironment for cancer cell growth and spread, and activation of carcinogenic signaling pathways^52^. Elevated LDH levels are a marker of cancer cells turnover rate, and LDH has a potential role for prediction of potential invisible metastases^44^. We found that all these markers had non-trivial on-treatment kinetics. However, data fits were not perfect, possibly due to the simplicity and empiric nature of the models we used. Further mechanistic modeling of the joint kinetics of BKs and TK could bring relevant biological information and yield more accurate predictive parameters. We found that all four BKs were contributive to the model and that, combined, they outperformed TK performances.

We analyzed the RNAseq data using standard methods and found only negligible predictive performances. Such result could be explained by the fact that the tissue of origin that was used was heterogeneous across the patients (primary tumor or metastasis), was limited to a local area of the tumor, and could come from tissue sampled long before treatment initiation. Given that our main objective was to derive a predictive model from markers available in routine practice, we excluded it from our minimal signature. A refined analysis, especially focusing on immune-based signatures, could improve our results^5^.

Machine learning models, although increasingly used in pharmacological studies —including recently for TK-OS modeling and variable selection^22,53^ —have yet rarely been rigorously compared to classical statistical models^24^. Here, such comparison revealed significantly better performance of the nonlinear random survival forest RSF model compared to the linear proportional hazards Cox model. In our approach, we did not use the propagation of standard statistical quantification of the parameters’ estimates uncertainty to evaluate the accuracy of the model predictions. Rather, we relied on the RSF-outputted individual survival curves to sample virtual individuals and compute prediction intervals.

A drawback of classical TK–OS studies is that they make use of the full observed kinetics to predict overall survival, which can lead to time-dependent covariate bias^54^ and limit their practical applicability at bedside. We used individual-truncated data sets and found that kML was already improving predictions over mBSL using data at 1.5 months, which corresponds to the first imaging assessment of the treatment effect. At later times, stratification abilities increased to highly significant levels (e.g., *HR* = 5 at 6.8 months).

A strength of our study is that we relied on well-curated data with high number of patients from clinical trials. However, when extrapolating to other settings — earlier trial phases, real-world data — limited number of patients might be available. Yet, we found that using only 60 patients to train kML was sufficient to reach near-optimal performances.

Not only kML has value for personalized health care, but it also revealed useful for prediction of a phase 3 trial using early on-study data. Our model was able to predict the study’s positive outcome with data at ∼ 6 months, versus 10 months using TK only^19^. Relying on the data alone, such positive outcome was only detectable at 15 months. These results could have important implications for drug development as they could inform earlier on go/no-go decisions. Consequently, this could allow to detect futility more easily and more rapidly during clinical trials, allowing to avoid treating patients with an inefficient investigational treatment and to reassign funds and energy to other researches. Of note, in a recent evaluation based on resampling the first-line NSCLC ATZ study IMpower150 to mimic small, short follow up early Phase Ib studies, TK model-based metrics had better operating characteristics to predict Phase III success compared with RECIST endpoints ORR and PFS^55^. Extension of such results with the addition of BKs is thus a promising line of research. In addition, kML, trained on ATZ data, yielded excellent predictive abilities for the docetaxel (control) arm. This suggests that the relationships between TK / BK and OS might be drug-independent. In turn, this opens future perspectives in terms of testing kML on drugs with different mechanism of action, or combinations.

Further avenues of research comprise the development of integrative models from advanced multi-modal data such as the one collected during the PIONeeR clinical study (NCT03833440)^56,57^ that include quantitative image analysis from multiplex immune–histochemistry, genomic and transcriptomic data, biological and clinical markers. In addition, mechanistic modeling of quantitative and physiologically meaningful longitudinal data (immune-monitoring, vasculo-monitoring, circulating DNA^12,58–60^, soluble factors^61^, pharmacokinetics, TK and a large number of BKs from either hematology or biochemistry) paves the way to an improved understanding and prediction of mechanisms of relapse to ICI^62^. Furthermore, the predictive abilities of kML — at the both individual and study levels — should be evaluated in model-based prospective trials^63^.

In conclusion, our study shows that integrating model-based on-treatment dynamic data from routine biological markers shows great promise for both personalized health care and early prediction of the outcome of clinical trials during drug development.

## Data Availability

Qualified researchers may request access to individual patient level data through the clinical study data request platform (https://vivli.org/). Further details on Roche's criteria for eligible studies are available here (https://vivli.org/members/ourmembers/). For further details on Roche's Global Policy on the Sharing of Clinical Information and how to request access to related clinical study documents please refer to the Roche website (https://www.roche.com/innovation/process/clinical-trials/data-sharing/).

https://www.roche.com/innovation/process/clinical-trials/data-sharing/

## Supplementary Figures

**Supplementary Figure 1.**
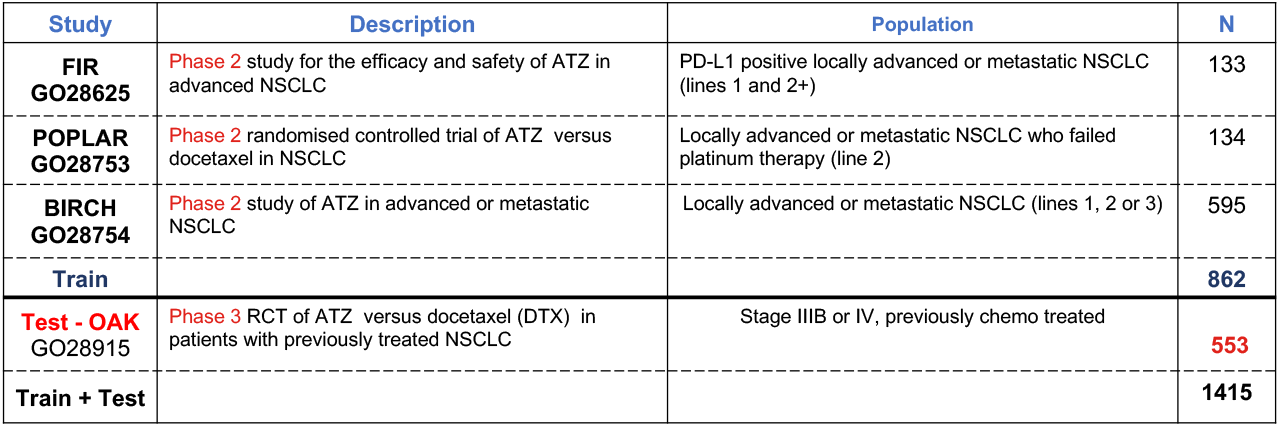
Train and test data sets Four monotherapy studies of atezolizumab in advanced NSCLC. NSCLC: Non-Small Cell Lung Cancer; p = number of parameters, N: number of patients treated with atezolizumab (patients from French centers were excluded for legal reasons (N=118); In total, data from 1074 patients from OAK were used as Test set (553 from the ATZ arm, 521 from the DTX arm); PD: Pharmacodynamic; SLD: Sum of the Largest Diameters. CRP: C Reactive Protein; LDH: Lactate Dehydrogenase.

**Supplementary Figure 2.**
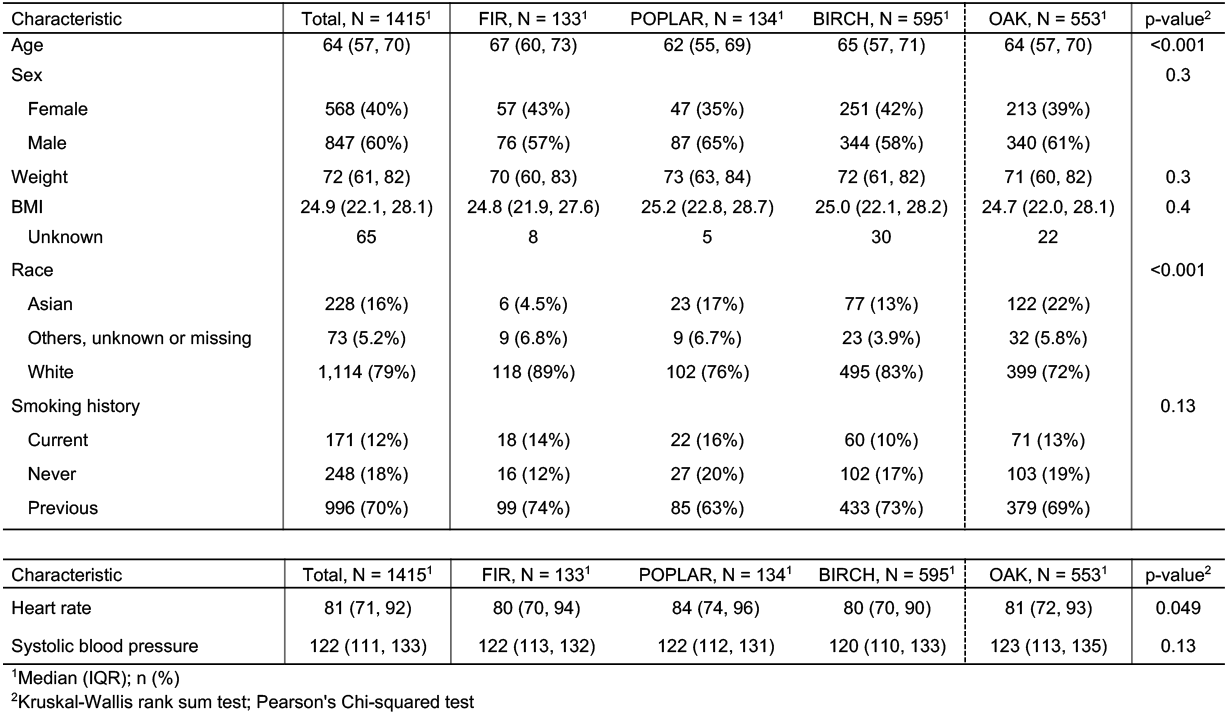
Patient characteristics: demographics and clinics

**Supplementary Figure 3.**
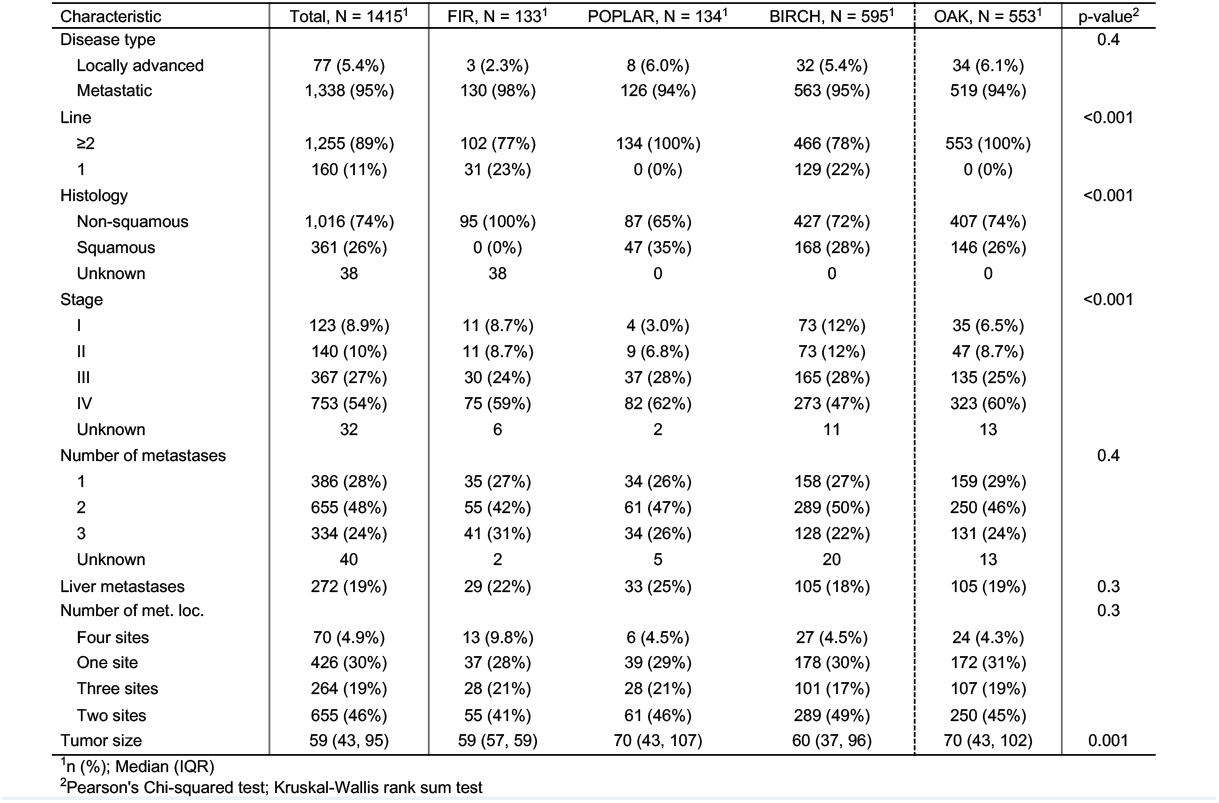
Patient characteristics: disease

**Supplementary Figure 4.**
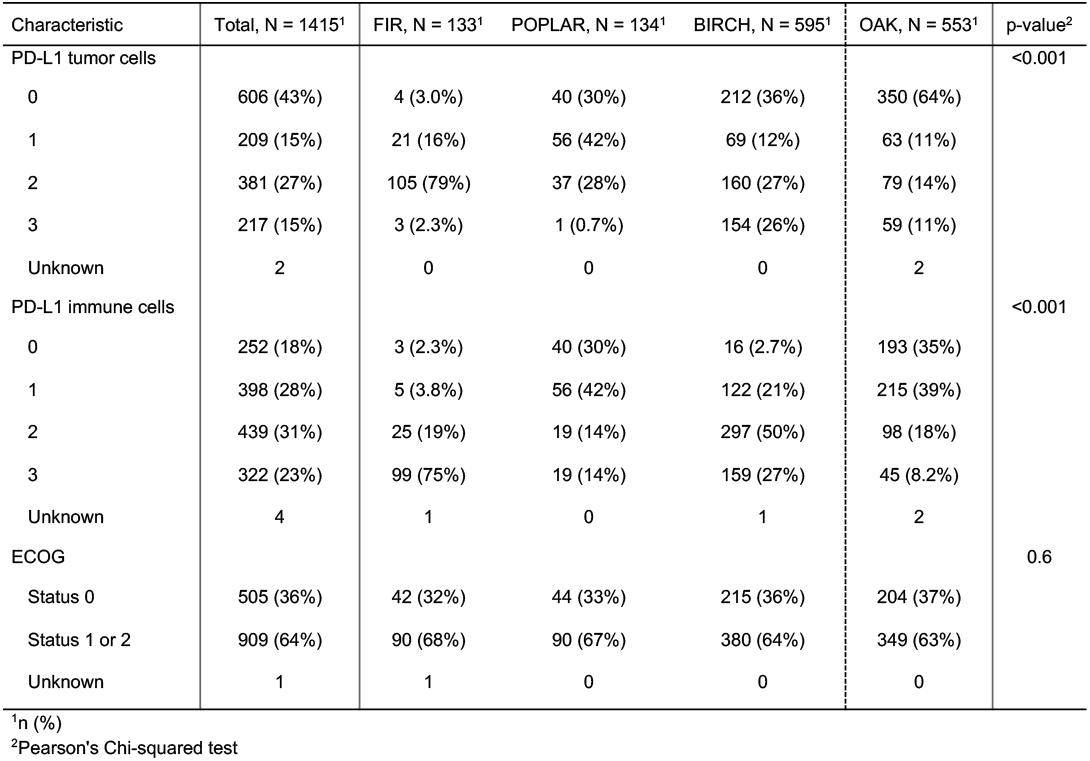
Patient characteristics: PD-L1 and ECOG

**Supplementary Figure 5.**
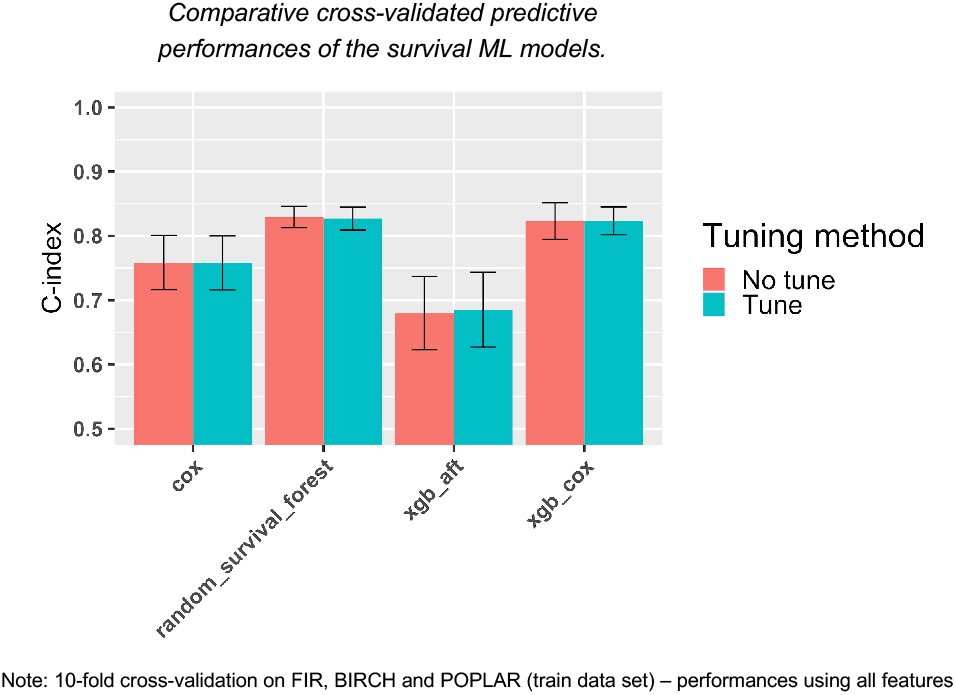
Comparison of ML algorithms and tuning methods

**Supplementary Figure 6.**
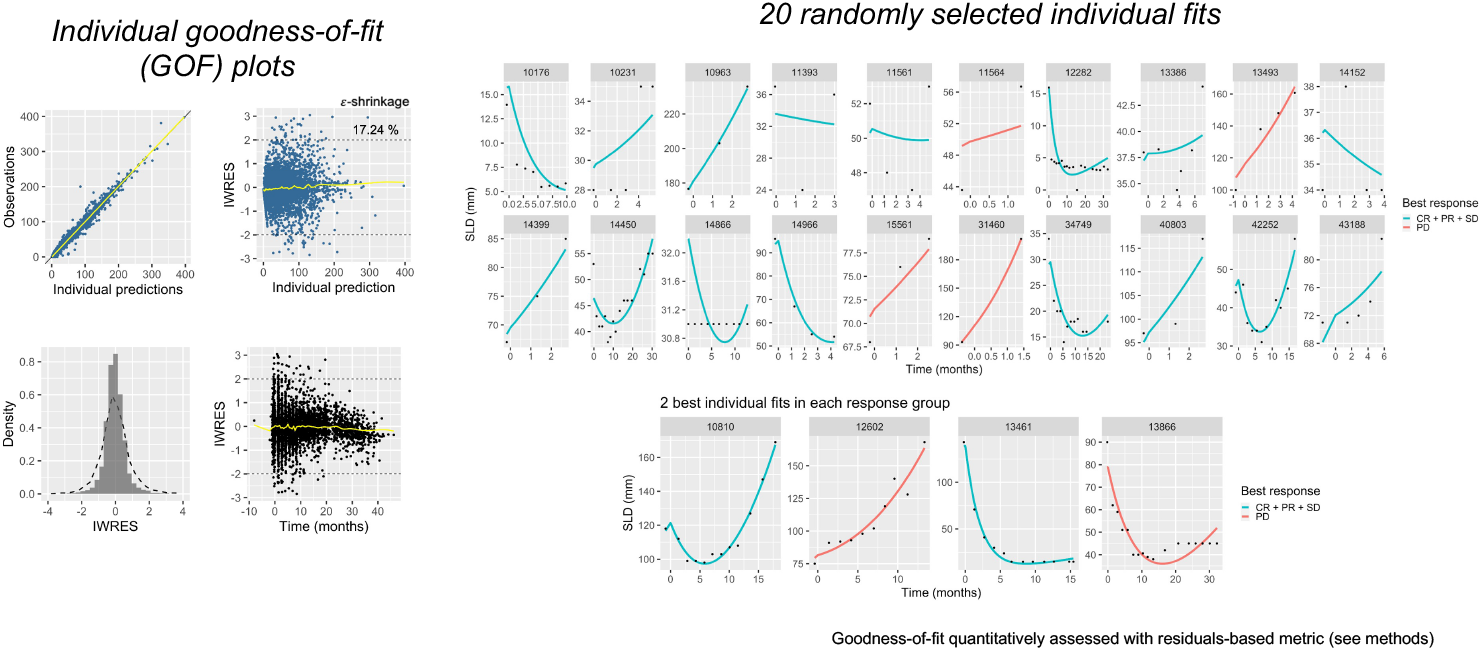
TK modeling goodness-of-fit

**Supplementary Figure 7.**
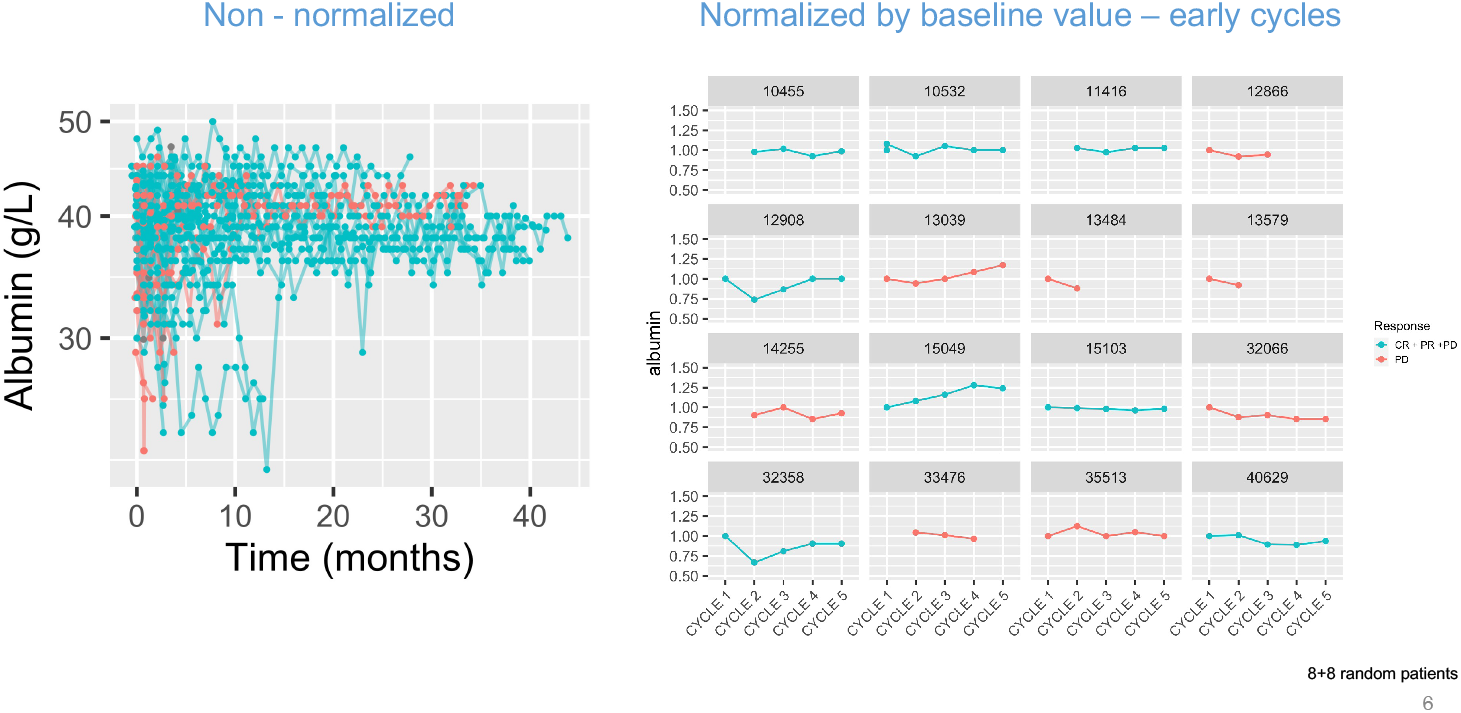
Examples of longitudinal kinetics: Albumin

**Supplementary Figure 8.**
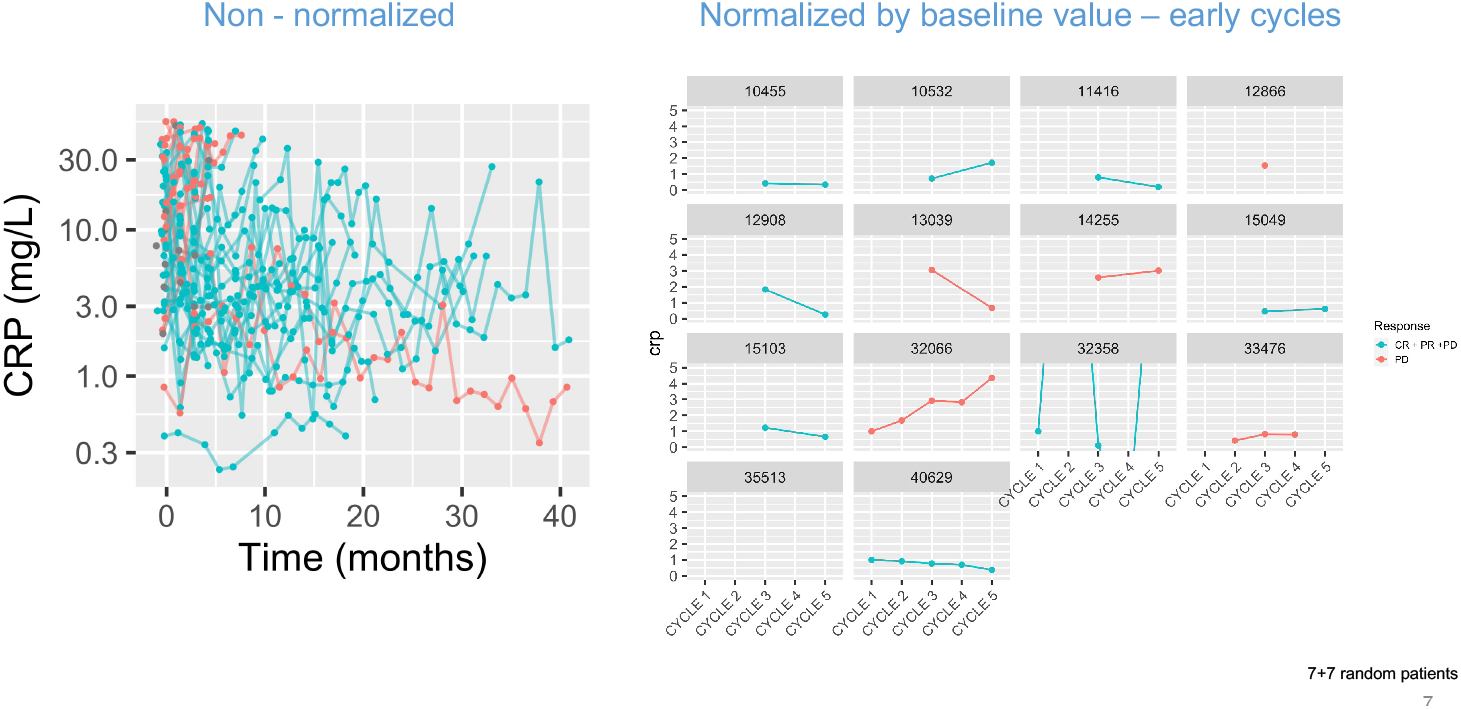
Examples of longitudinal kinetics: CRP

**Supplementary Figure 9.**
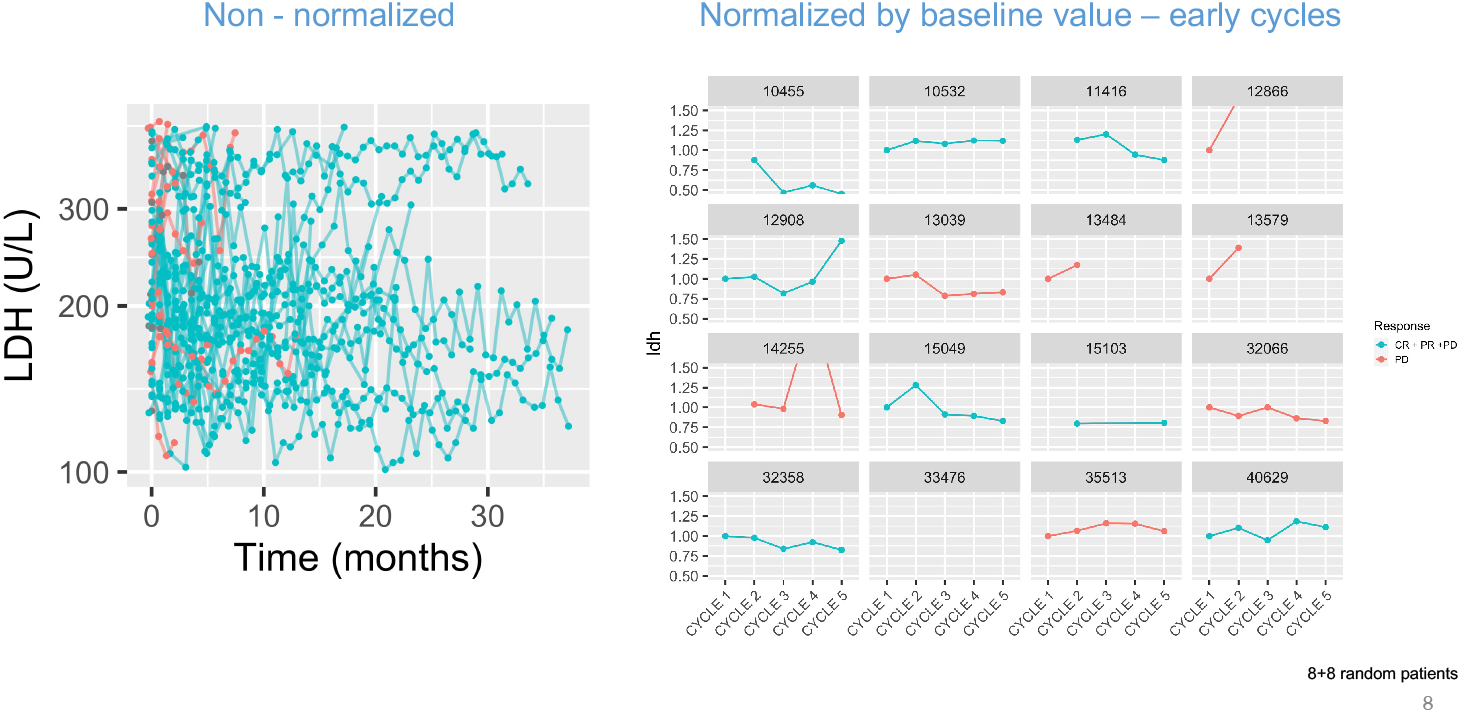
Examples of longitudinal kinetics: LDH

**Supplementary Figure 10.**
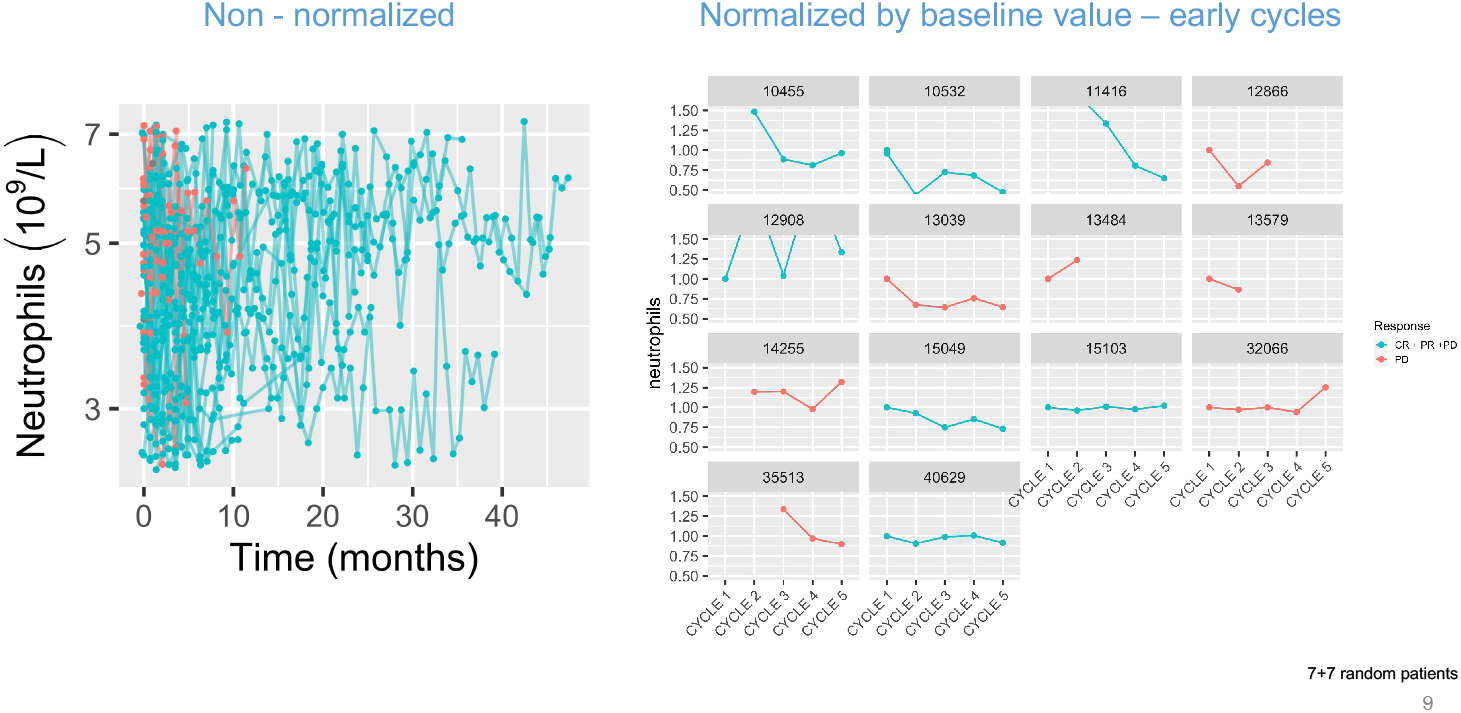
Examples of longitudinal kinetics: Neutrophils

**Supplementary Figure 11.**
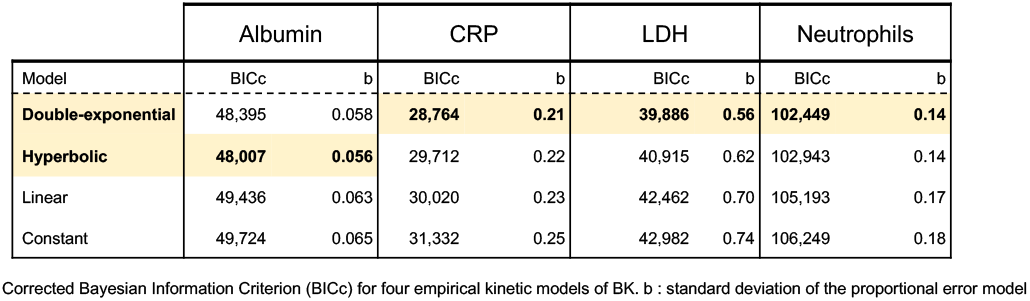
Goodness-of-fit metrics of dynamic BK models

**Supplementary Figure 12.**
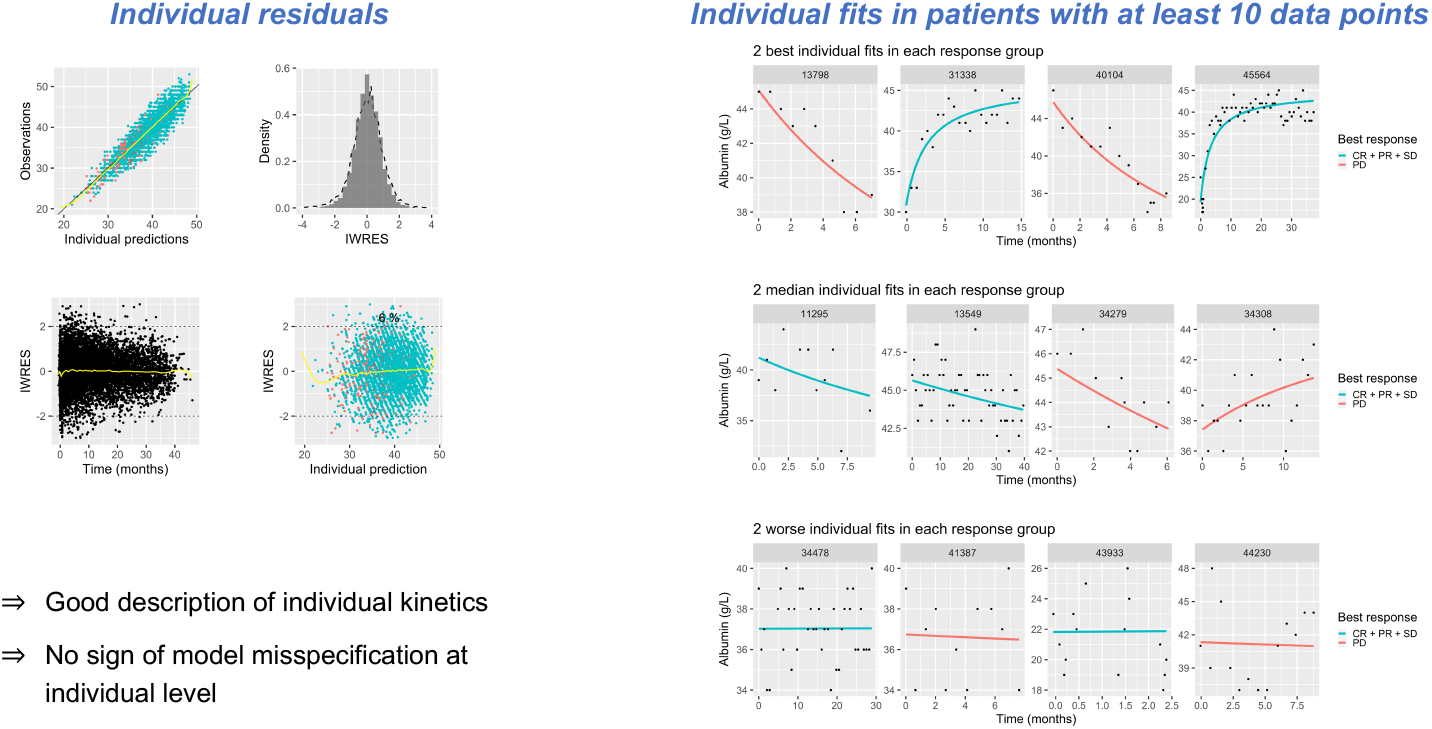
Albumin: hyperbolic individual fits

**Supplementary Figure 13.**
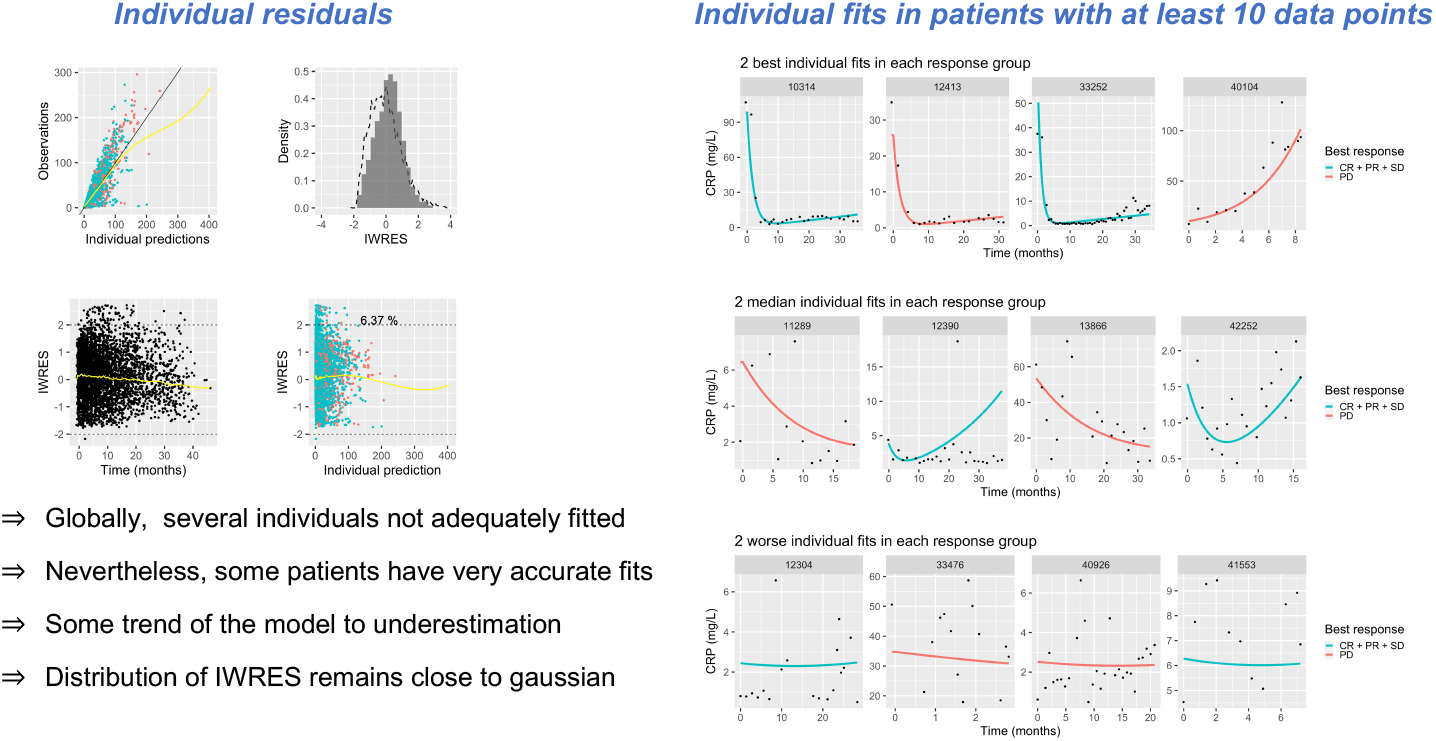
CRP: dexp individual fits

**Supplementary Figure 14.**
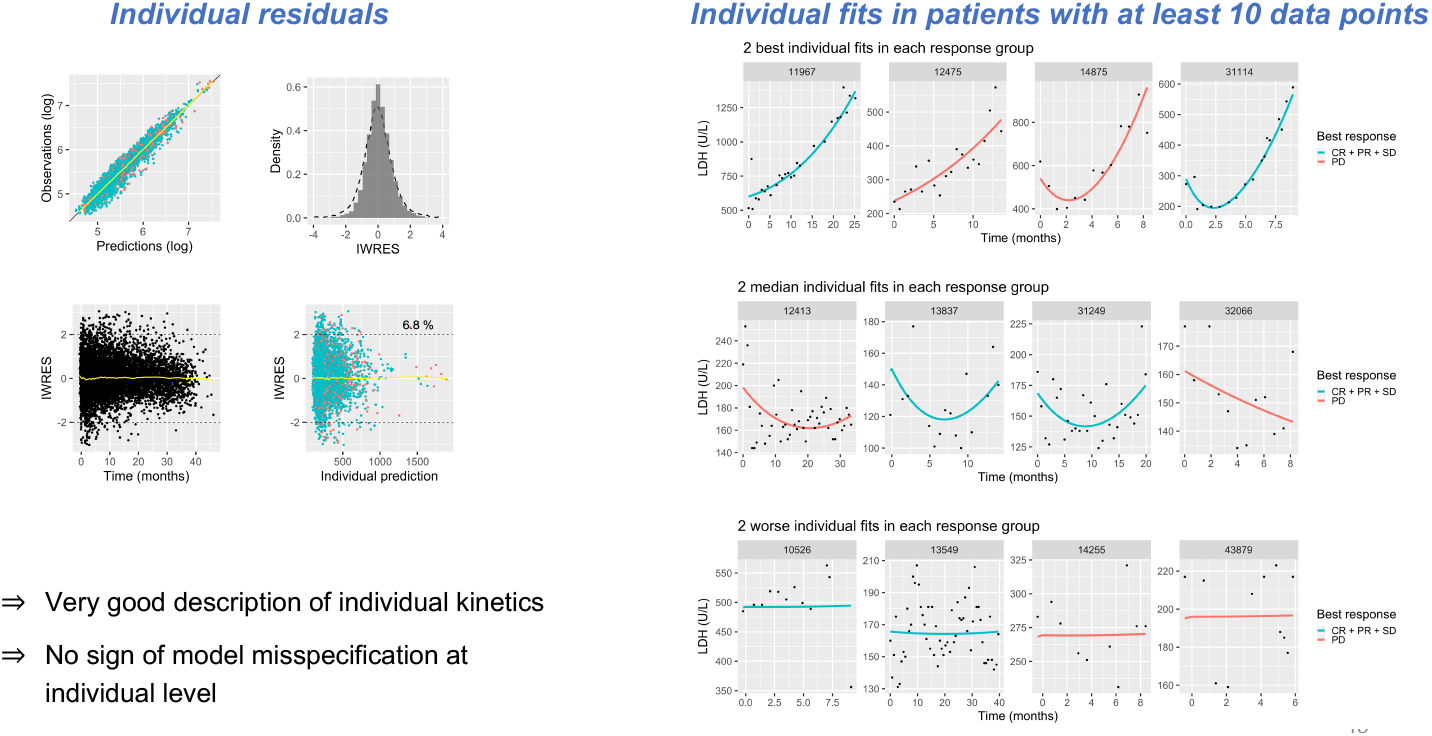
LDH: dexp individual fits

**Supplementary Figure 15.**
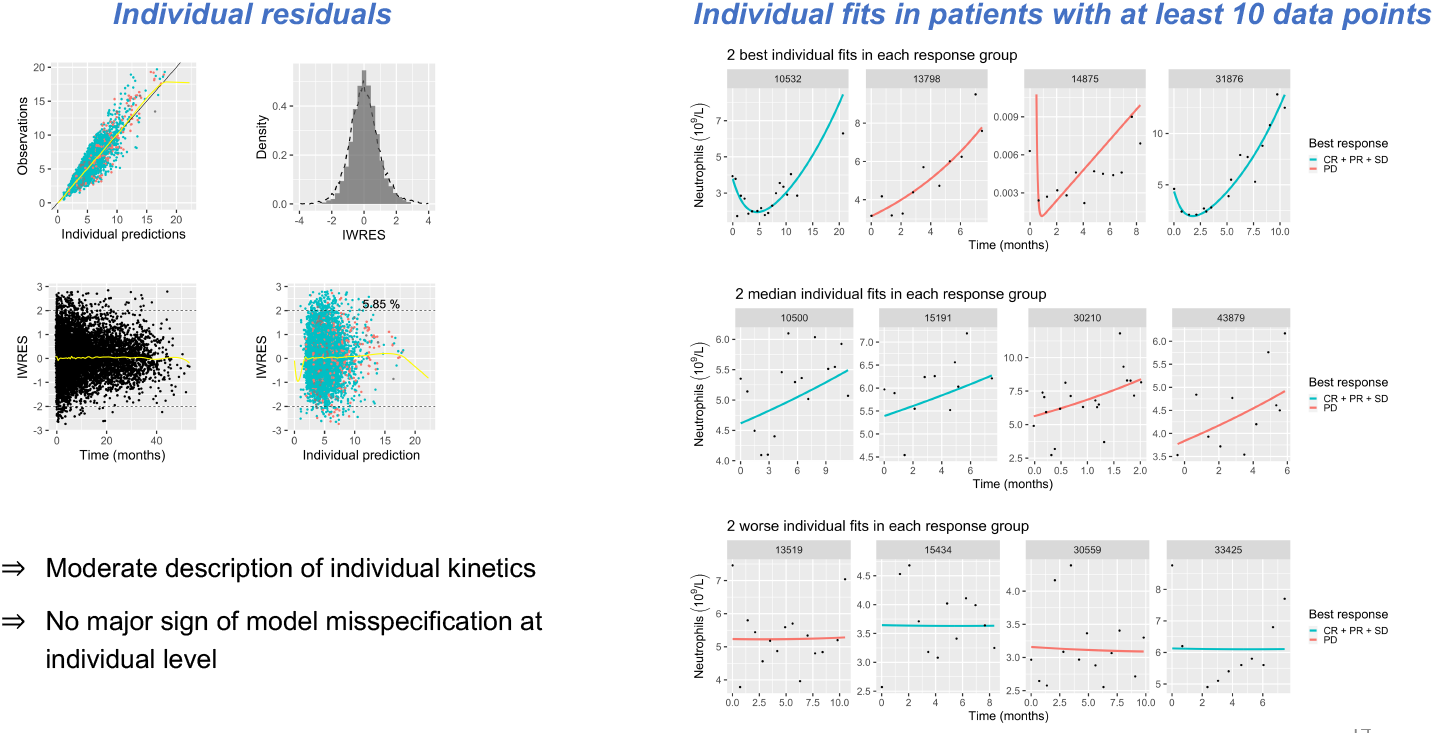
Neutrophils: dexp individual fits

**Supplementary Figure 16.**
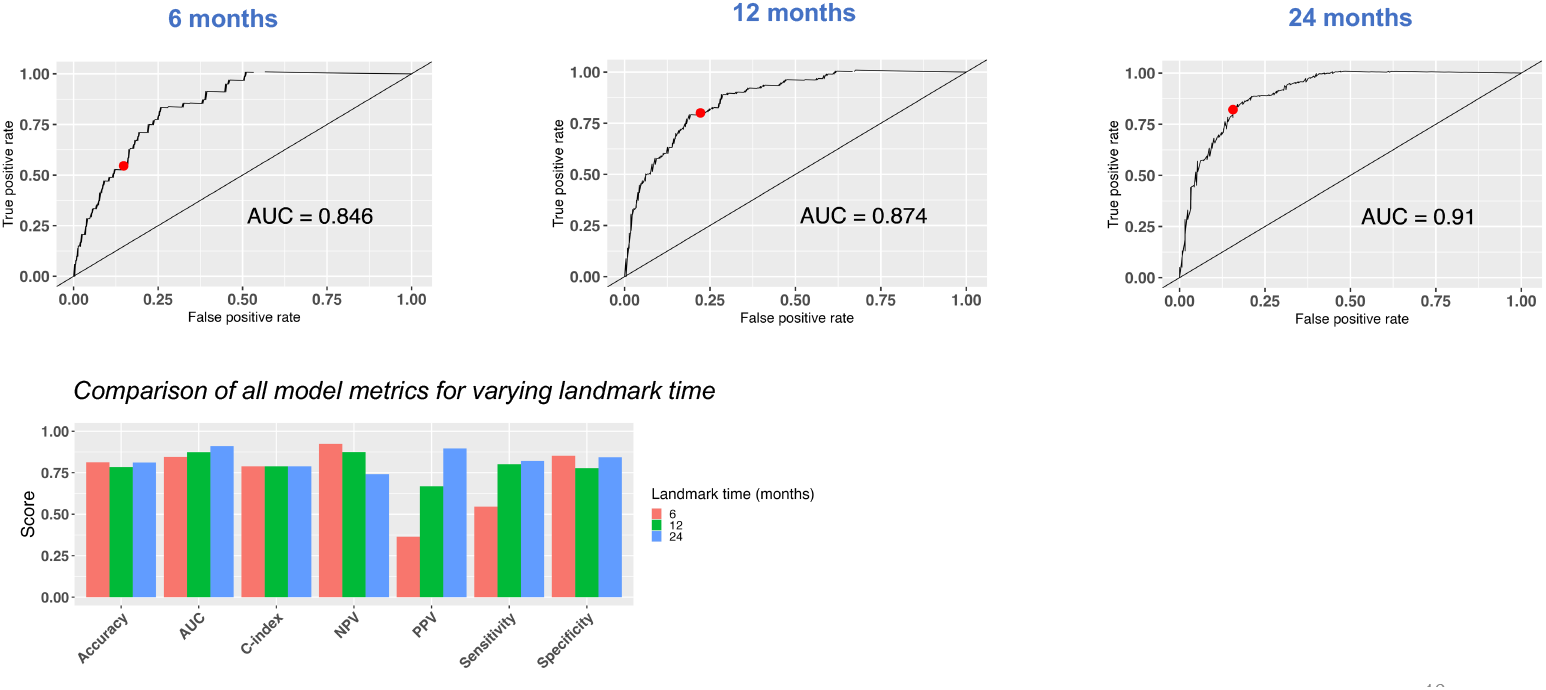
ROC curves for variables landmark times (test set - OAK)

**Supplementary Figure 17.**
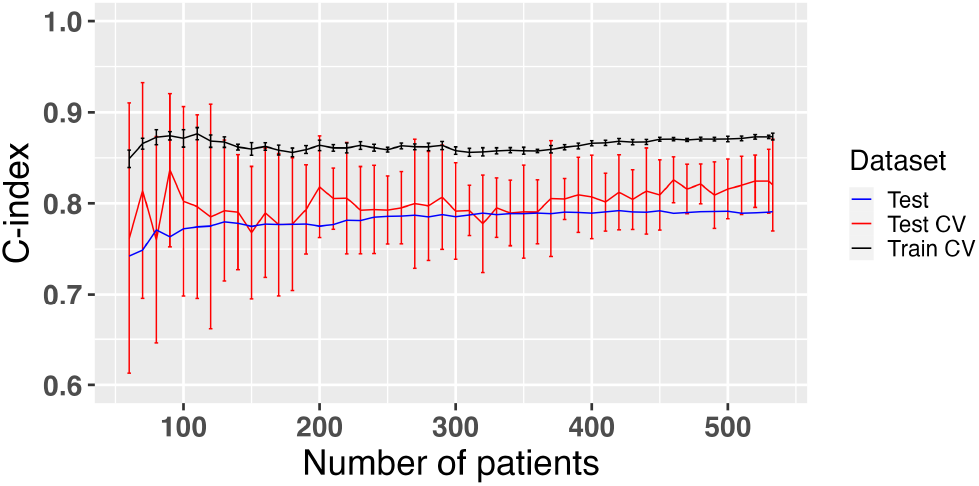
Learning curve

**Supplementary Figure 18.**
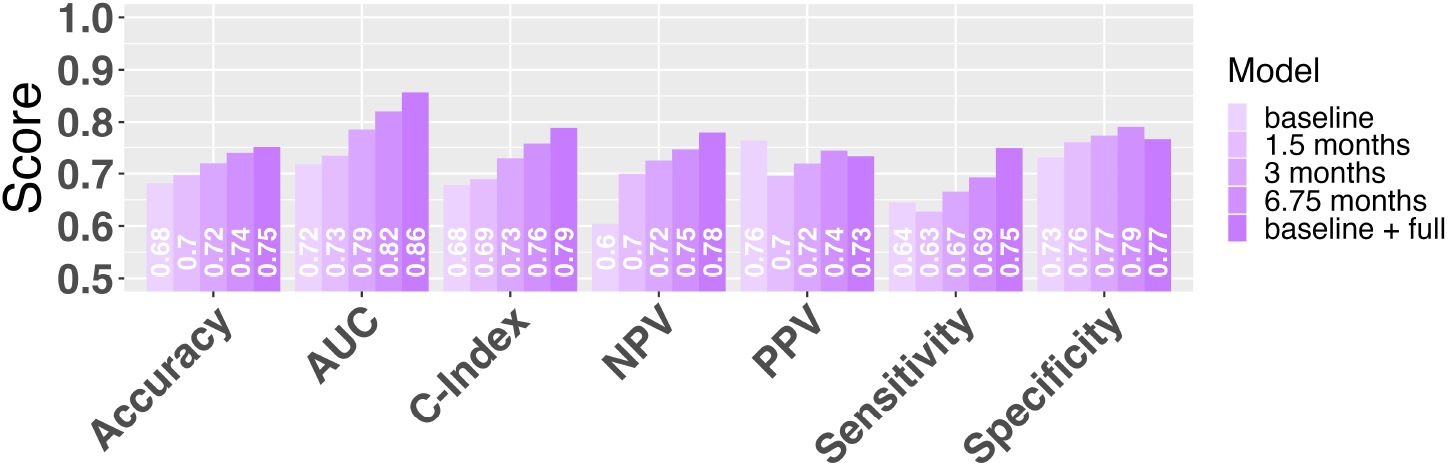
Additional value of NLME to baseline for multiple metrics

**Supplementary Figure 19.**
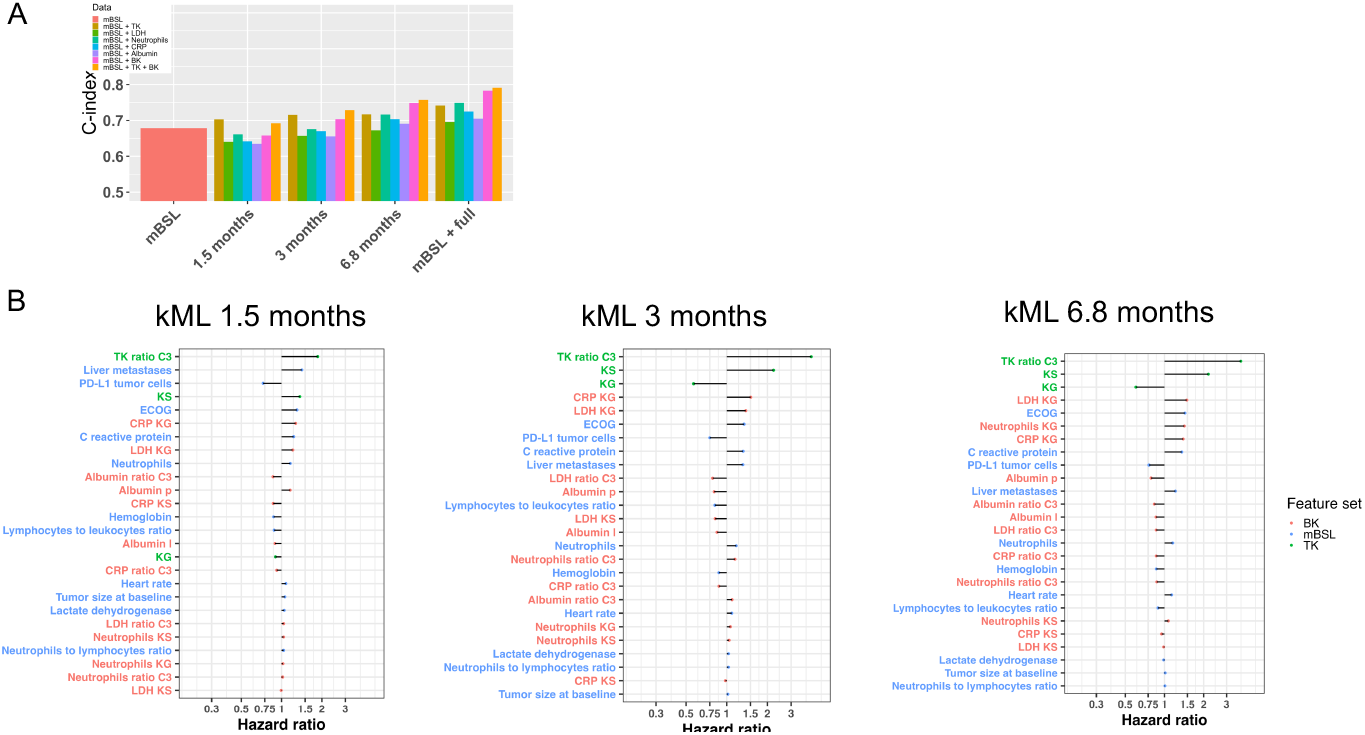
kML models using only single kinetic markers

## Supplementary Methods

### Dimensionality reduction for RNAseq

Initial expression data from RNAseq consisted of 715 patients and 58, 311 transcripts. The first step of data filtering removed all transcripts with less than 10 read counts for all patients, then selected genes with highest variability between patients (top 15, 000 transcripts most variable). Then, data were normalized using upper quartile normalization which consisted in dividing each read count by the 75^th^ percentile of the read counts of the corresponding sample and the final expression values were log_2_ transformed. Subsequently, a univariable Cox regression model was employed to statistically assess the correlations between the expression levels of the transcripts and overall survival. Bonferroni correction was used to adjust p-values from multiple univariate tests. This step was performed using the `RegParallel` R package. We selected transcripts with high predictive values using following criteria: adjusted log rank < 0.01 and HR < 0.85 or HR > 1.2. The remaining transcripts were used to perform a bootstrap Lasso Cox regression with cross-validation using mainly the `glmnet` R package. Finally, the smallest number of transcripts with best predictive model (highest C-index) was selected for further analysis.

### Rules for BK processing

1. Observations outside lower (LB) and upper (UB) physiological bounds were discarded using the following values, determined from discussion with a clinical oncologist: albumin, LB = 10 g L^−1^, UB = 100 g L^−1^; CRP, no LB, UB = 300 mg L^−1^; LDH, LB = 50 U/L, UB = 2000 U/L; neutrophils, no LB, UB = 20.
2. For duplicates, the first one recorded was kept.
3. Denoting BK_*n*_ the value of the a “BK” at time *t*_*n*_ for a given patient, we excluded values such that: BK_*n*_ ∉ (BK_*n-1*_, BK_*n+1*_) AND |BK_*n*_ − BK_*n-1*_| > 3 × *sd*_*BK*_ AND |BK_*n*_ − BK_*n+1*_| > 3 × *sd*_*BK*_, where *sd*_*BK*_ is the standard deviation of {BK_*n*_}_*n*_, i.e. all time points for this patient.
4. The BK value at the closest time point to treatment initiation was kept, provided this time point was no more than 40 days before or 10 days after treatment initiation (otherwise, patient was disregarded).

### Nonlinear mixed-effects modeling

Denoting by ℳ (*t*; *θ*) a structural dynamic model that depends on time *t* and a set of parameters *θ*, longitudinal observations 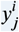 in patient *i* at time 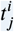 were assumed to follow the observation model

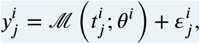

where 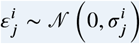 is the gaussian-distributed error model. The latter was either constant 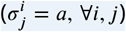 for TK or proportional 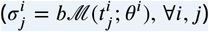 for BK. To describe inter-individual variability, individual parameters *θ*^i^ were assumed to follow log-normal distributions:

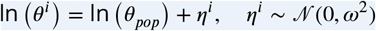

with population-level parameters *θ*_*pop*_ and *ω*. Estimation of these was performed using the stochastic approximation of expectation maximization algorithm implemented in the Monolix software.

